# Distinct Clinical Associations of Blood Tau Biomarkers and Neurofilament Light in Amyotrophic Lateral Sclerosis

**DOI:** 10.64898/2026.07.03.26356752

**Authors:** Bernat Bertran-Recasens, Paula Ortiz-Romero, Ferran Lugo-Hernández, Sergio Vidal Notari, Marina De Diego-Osaba, Helena Blasco-Forniés, Esther Jiménez-Moyano, Mireia Llop Trujillano, Javier Torres-Torronteras, Marta del Campo, Miguel Ángel Rubio Pérez, Marc Suárez-Calvet

## Abstract

**Background and Objectives:** To investigate the associations of blood-based tau biomarkers with clinical, electrophysiologic and prognostic measures in amyotrophic lateral sclerosis (ALS), and to determine whether they reflect distinct disease-related processes.

**Methods:** We studied 119 patients with ALS from a longitudinal observational cohort. Plasma and serum p-tau181, p-tau217, p-tau231, brain-derived tau (BD-tau), NfL and GFAP were measured using Lumipulse and Simoa assays. Associations with demographic variables, disease severity (ALSFRS-R and slow vital capacity), lower motor neuron (LMN), muscle involvement (creatine kinase [CK] and high-sensitivity cardiac troponin T [hs-cTnT]), disease progression and survival were assessed using multivariable models.

**Results:** Tau-related biomarkers, specifically p-tau217 and BD-tau, were associated with greater cross-sectional disease severity, reflected by lower ALSFRS-R scores. Plasma and serum p-tau181, p-tau217, p-tau231, and BD-tau were associated with higher CK and hs-cTnT, whereas p-tau181 and p-tau231 were also associated with greater LMN involvement. In contrast, NfL and GFAP were not associated with muscle or LMN involvement. Across analytical platforms, plasma and serum NfL were associated with faster ALSFRS-R decline and shorter survival. NfL was the only biomarker independently associated with both disease progression and survival.

**Discussion:** Blood biomarkers capture distinct dimensions of ALS. Tau-related biomarkers are associated with cross-sectional disease severity, LMN involvement and muscle injury, whereas NfL primarily reflects disease progression and survival. These findings support the complementary use of tau-related biomarkers and NfL for ALS phenotypic characterization and prognosis assessment.

## Introduction

Amyotrophic lateral sclerosis (ALS) is a progressive neurodegenerative disorder characterized by degeneration of upper (UMN) and lower motor neurons (LMN), leading to weakness, disability, respiratory failure and death, typically within 2–5 years of symptom onset.^1^ ALS is clinically heterogeneous, with variability in site of onset, burden of upper versus lower motor neuron involvement, progression rate and survival.^2^ This heterogeneity has direct implications for diagnosis, prognosis, patient counseling, patient stratification and clinical trial design.^3^

Blood-based biomarkers are increasingly used in neurodegenerative diseases, contributing to diagnostic prognostication, disease monitoring and clinical trial enrichment.^4^ These biomarkers are widely developed in Alzheimer’s disease (AD) and multiple sclerosis,^5–7^ where they have contributed to the development of disease-modifying therapies.^8^ A similar potential exists in ALS, as evidenced by the development of antisense oligonucleotide (ASO) therapies for SOD1-and FUS-related forms of the disease.^9–11^

In ALS, neurofilament light chain (NfL) is the most established blood biomarker and has been consistently validated as a marker of neuronal and axonal injury, disease progression and survival.^12–17^ Although NfL provides important prognostic information, it may be less informative regarding other clinically relevant disease dimensions, such as cross-sectional disease severity and the anatomical distribution of disease burden, including the relative contribution of UMN and LMN involvement.^13,18,19^ This creates an important biomarker gap in ALS: beyond predicting progression and survival, there is a need for blood biomarkers that capture clinically relevant disease dimensions. Several candidate biomarkers originally developed in AD research, including phosphorylated tau (p-tau)181, p-tau217 and p-tau231 and glial fibrillary acidic protein (GFAP) have garnered growing attention.^20–25^ Elevated blood levels of p-tau181 and p-tau217 have been reported in ALS, raising the possibility that blood tau elevations in this disease may be partly attributable to peripheral sources.^26–35^ In parallel, biomarkers reflecting muscle and LMN involvement, such as high-sensitivity cardiac troponin T (hs-cTnT), have also emerged as promising tools for ALS phenotyping and disease characterization.^18,36,37^

Recent studies have further supported this concept. Blood p-tau181 and p-tau217 have been shown to be increased in ALS and detectable in skeletal muscle, with increased p-tau immunoreactivity in atrophic muscle fibers.^29^ These findings suggest that blood tau biomarkers in ALS may reflect LMN denervation or muscle-related pathology rather than AD-type central tau pathology alone.^29,30^ However, prior studies have mainly focused on selected tau species, usually p-tau181 or p-tau217, and have not systematically compared multiple tau-related biomarkers across plasma and serum against detailed clinical measures of LMN involvement, muscle injury, disease severity, progression and survival.^27,38–40^

BD-tau is a newer blood biomarker designed to preferentially measure brain-derived tau species and has shown promise as a marker of AD-related neurodegeneration.^41^ Its relevance in ALS remains uncertain. Whether BD-tau reflects central neurodegeneration, LMN involvement, muscle pathology or other disease processes in ALS is currently unclear.

We therefore aimed to determine whether blood-based tau biomarkers (p-tau181, p-tau217, p-tau231, BD-tau), NfL and GFAP are differentially associated with disease severity, LMN and muscle involvement, disease progression and survival in ALS.

## Methods

### Study design and participants

This was a single-center, longitudinal observational cohort study. We included consecutive participants who met the revised El Escorial criteria^42^ for definite or probable ALS between February 10, 2020, and July 15, 2025, at Hospital del Mar, Barcelona, Spain (n = 119).

### Clinical Assessments

At the study visit, blood samples were obtained for routine clinical analyses, including creatine kinase (CK) and creatinine, and additional blood samples were collected for biomarkers’ measurements (see sample processing and biomarker measurements). Neurologists specialized in ALS obtained medical history, conducted neurological examinations and reviewed diagnostic investigations. The following demographic and clinical variables were determined: age, symptom of onset, body mass index (BMI), in kg/m^2^, disease classification according to the revised El Escorial criteria (definite or probable),^42^ the Revised ALS Functional Rating Scale (ALSFRS-R),^43^ King’s stage as disease staging system,^44^ and riluzole treatment status. In addition, respiratory function was assessed using slow vital capacity (SVC), expressed as percentage of the predicted value,^45^ and the extent of LMN involvement was evaluated.

The assessment of LMN involvement was conducted according to the methodology described by Chamoun et al.^18^ Participants were included in this analysis if their neurophysiological evaluation had been performed within the 6 months prior to sample collection and included muscles from at least three of the four anatomical regions. A region was considered affected when the electromyography (EMG) study demonstrated active fibrillation potentials and positive sharp waves in at least one muscle. For each participant with EMG available, we assessed the number of LMN-affected regions (0–4) as well as the proportion of affected muscles. The proportion of affected muscles was calculated relative to the total number of muscles examined and expressed as a percentage using the following formula: (number of affected muscles / total number of muscles examined) × 100 (range: 0–100%).

Disease duration was defined as the time in months between symptom onset and blood sampling. Disease progression rate (ALSFRS-R slope) was calculated as (48 − ALSFRS-R score at blood sampling) / disease duration in months.

Genetic testing results were reviewed when available. Pathogenic mutations were identified in *FUS* (n = 5), *SOD1* (n = 1), *TBK1* (n = 2), and *ARPP21* (n = 2) genes, as well as *C9orf72* hexanucleotide repeat expansions (n = 7) and an *ATXN2* expansion (n = 1). No pathogenic mutations were identified in 59 patients, while 42 patients did not undergo genetic assessment. Genetic testing was not performed uniformly across the cohort and reflected evolving clinical practice and patient preference.^46^ Between 2020 and 2022, genetic testing (*C9orf72* expansion and *SOD1*) was only performed in patients younger than 45 years, with a family history of ALS, or with FTD/cognitive/behavioural features. From 2023 onwards, *C9orf72* expansion testing plus exome sequencing was proposed to all patients regardless of age, phenotype or family history.

### Sample processing and biomarker measurements

Blood samples were obtained during the ALS clinic visit. Serum samples were collected in BD Vacutainer® tubes (Cat. No. 368815) with a serum separator and allowed to clot upright at room temperature for 1–2 hours. After centrifugation at 1800 × g for 10 minutes at 4 °C, the serum was transferred into sterile cryogenic conical microtubes (Thermo Fisher Scientific Inc. Cat. No. 377267) and stored at −80 °C. Plasma samples were collected in BD Vacutainer® K2E EDTA tubes (Cat No. 367525), centrifuged at 1800 × g for 10 minutes at 4 °C within 2 hours of collection, aliquoted and stored at −80 °C.

Plasma and serum NfL were measured using 3 assays: NfL Lumipulse (Lumipulse G NFL Blood, Fujirebio), NfL (singleplex) Simoa (NF-Light Advantage PLUS, Quanterix) and NfL (N4PD) Simoa (Neurology 4-plex D Advantage PLUS, Quanterix). Tau-related biomarkers were measured using the p-tau217 Lumipulse (Lumipulse G p-tau 217 Plasma, Fujirebio) and p-tau181 Simoa (pTau 181 Advantage PLUS, Quanterix), and Neurology 4-plex D Advantage PLUS assay was used for measuring brain-derived tau (BD-Tau) and glial fibrillary acidic protein (GFAP). Plasma p-tau231 was measured using the in-house assay developed at the University of Gothenburg (UGot p-tau231).^22,47^

All these biomarkers were measured at the Fluid Biomarkers Facility of the BBRC (Barcelona, Spain). Fujirebio assays were performed on a Lumipulse G1200 analyzer. Samples were analyzed in singlicates. Signal variations within and between analytical runs were assessed using an internal Quality Control (iQC) plasma sample that was run every 50 samples. Quanterix assays were performed on a Simoa HD-X analyzer. Samples were analyzed in duplicates for the NF-Light and in singlicates for the remaining Quanterix assays. Signal variations within and between analytical runs were assessed using two iQC plasma samples analyzed in duplicate at the beginning and the end of each plate. Intra- and inter-assay % coefficient of variation (CV) derived from the iQC samples are reported in eTable 1.

CK and hs-cTnT were measured at Hospital del Mar using the fifth-generation Elecsys assay and direct enzymatic method (Roche Diagnostics), respectively.

Laboratory personnel were blinded to clinical data.

### Ethics

The study was approved by the Hospital del Mar Clinical Research Ethics Committee (2019/8803/I) and followed the ethical principles of the Declaration of Helsinki. All included patients gave their written informed consent to participation prior to their inclusion in the study.

### Statistical analyses

Participant characteristics were summarized as median (IQR) for continuous variables and frequency (n) and percentage (%) for categorical variables. Comparisons of non-biomarker variables between groups were performed according to variable type and distribution. Continuous variables were analyzed using the Wilcoxon rank-sum test, whereas categorical variables were analyzed using Pearson χ² tests. Missing data was present, and the number of available observations is reported in the tables. All analyses were performed using complete-case analysis.

Correlations between non-transformed biomarker concentrations and continuous demographic variables, as well as correlations among non-transformed biomarker concentrations, were assessed using Spearman’s rank correlation coefficient.

For all subsequent analyses, biomarker concentrations were natural log-transformed prior to analysis to reduce skewness and improve model fit. The distribution of biomarker values was assessed using density plots, Q–Q plots, boxplots and formal normality tests (Shapiro–Wilk and/or Lilliefors tests). For survival analyses, biomarker values were additionally standardized as z-scores using the whole cohort as the reference group, allowing direct comparison of effect estimates across blood-based biomarkers measured on different analytical platforms.

For group comparisons, log-transformed biomarker concentrations were modeled as dependent variables in ANCOVA models. The independent variable of interest varied according to the comparison performed (sex, onset form or riluzole treatment). Three main models were evaluated: Model 1 (unadjusted), Model 2 (adjusted for demographic and biological covariates potentially influencing biomarker values: age, sex, BMI, estimated glomerular filtration rate [eGFR]) and Model 3 (additionally adjusted for ALS clinical variables, including onset form, King’s stage, riluzole treatment and ALSFRS-R score). An exploratory sensitivity model (Model 4), additionally adjusted for the number of LMN-affected regions, was performed to assess whether biomarker associations were independent of LMN burden. Because LMN involvement was derived from EMG data, Model 4 was restricted to participants with available neurophysiological assessments (n =67) and was conducted only for selected analysis. The adjustment strategy was adapted to each outcome to avoid adjustment for the outcome variable itself. P-values were corrected for multiple testing using the Benjamini–Hochberg FDR procedure for Models 3 and 4. Detailed model specifications are provided in the corresponding tables.

Linear regression models were used to assess the associations between log-transformed biomarker levels and continuous variables, including ALSFRS-R, SVC, CK, hs-cTnT, number of LMN-affected regions and proportion of affected muscles. The same hierarchical adjustment strategy (Models 1–4 described above) was applied across regression analyses, depending on the specific outcome and hypothesis being evaluated. P-values were also corrected for multiple testing using the Benjamini–Hochberg FDR procedure for Models 3 and 4. Model assumptions were assessed through diagnostic plots.

Survival analyses were performed using Cox proportional hazards regression models. All included participants were followed up until 15 July 2025. By this date, each individual had either experienced the event of interest or was classified as censored. Censoring was defined as being alive or having died from causes unrelated to ALS by 15 July 2025. The event was defined as ALS-related death or advanced respiratory failure (dependence on noninvasive mechanical ventilation for ≥22 hours per day or tracheostomy);^48^ none of the participants met the respiratory criteria. Patients carrying *FUS* pathogenic variants (n=5) were excluded from survival analyses because they were receiving antisense oligonucleotide treatment through an expanded access program, which could confound survival outcomes. However, blood sampling and clinical assessments were performed before treatment initiation. Hazard ratios (HRs) with 95% confidence intervals (CIs) were reported. The proportional hazards (PH) assumption was assessed for each Cox proportional hazards model using Schoenfeld residuals. Both biomarker-specific and global tests were examined; and a two-sided p-value <0.05 was considered evidence of a potential violation of the PH assumption. P-values were corrected for multiple testing using the Benjamini–Hochberg FDR procedure for Models 3 and 4.

In all cases two-sided p-values were reported, and statistical significance was defined as p or q < 0.05. All analyses were performed using version 4.5.0 of R (https://www.r-project.org/).

We used the STROBE reporting guideline^49^ to draft this manuscript, and the STROBE reporting checklist^50^ when editing. The checklist is included in eTable 2.

### Data Availability

The data used in this study were obtained from Hospital del Mar (Barcelona). Anonymized data not published within this manuscript can be made available on request for research purposes.

## Results

### Demographics and blood-based biomarkers

Demographic and clinical characteristics of the 119 patients with ALS included in this study are summarized in Table 1. The median age was 64 years (IQR, 53–74), and 67 patients (56%) were women. Spinal onset was the most frequent presentation (67%), and 73 patients (61%) were treated with riluzole. At baseline, the median ALSFRS-R score was 32 (IQR, 24–40), median SVC was 74% (IQR, 59–90), and the median progression rate was 0.86 ALSFRS-R points per month (IQR, 0.48–1.50). During follow-up, 71 patients (60%) died.

**Table 1.** Demographic, clinical, and biomarker characteristics of the ALS cohort.

| Characteristic | N | ALS cohort |
| --- | --- | --- |
| **Demographics and general characteristics** |  |  |
| Age, years | 119 | 64 (53, 74) |
| Sex | 119 |  |
| Female |  | 67 (56%) |
| Male |  | 52 (44%) |
| BMI, kg/m<sup>2</sup> | 119 | 23.1 (19.8, 26.7) |
| eGFR, mL/min/1.73 m<sup>2</sup> | 117 | 104 (94, 114) |
| **Clinical phenotype** |  |  |
| ALS onset form | 119 |  |
| Bulbar |  | 39 (33%) |
| Spinal |  | 80 (67%) |
| Riluzole treatment | 119 | 73 (61%) |
| **Disease severity** |  |  |
| King's stage | 119 |  |
| Kings 1/2/3 |  | 53 (45%) |
| Kings 4 |  | 66 (55%) |
| ALSFRS-R, points | 119 | 32 (24, 40) |
| SVC, % predicted | 81 | 74 (59, 90) |
| **Disease progression and survival** |  |  |
| ALSFRS-R slope, points lost/month | 119 | 0.86 (0.48, 1.50) |
| Survival time from symptom onset, months | 119 | 32 (20, 45) |
| Death during follow-up | 119 | 71 (60%) |
| **Lower motor neuron and muscle involvement** |  |  |
| EMG | 119 |  |
| Available |  | 67 (56%) |
| Not available |  | 52 (44%) |
| % Affected muscles | 63 | 76 (57, 88) |
| Number LMN-affected regions | 67 | 3 (2,3) |
| CK, U/L | 90 | 126 (65, 254) |
| hs-cTnT, ng/L | 113 | 23 (14, 35) |
| **Blood-based Biomarkers** |  |  |
| Plasma NfL (Lumipulse), pg/mL | 119 | 95 (60, 154) |
| Serum NfL (Lumipulse), pg/mL | 119 | 96 (64, 151) |
| Plasma NfL (singleplex, Simoa), pg/mL | 119 | 62 (40, 93) |
| Serum NfL (singleplex, Simoa), pg/mL | 119 | 84 (55, 119) |
| Plasma NfL (N4PD, Simoa), pg/mL | 119 | 64 (39, 91) |
| Serum NfL (N4PD, Simoa), pg/mL | 119 | 83 (51, 123) |
| Plasma p-tau181 (Simoa), pg/mL | 118 | 44 (28, 81) |
| Serum p-tau181 (Simoa), pg/mL | 116 | 35 (17, 59) |
| Plasma p-tau217 (Lumipulse), pg/mL | 119 | 0.19 (0.14, 0.35) |
| Serum p-tau217 (Lumipulse), pg/mL | 119 | 0.20 (0.14, 0.37) |
| Plasma p-tau231 (Simoa), pg/mL | 116 | 4.5 (2.7, 7.3) |
| Serum p-tau231 (Simoa), pg/mL | 114 | 1.75 (1.10, 3.12) |
| Plasma BD-tau (Simoa), pg/mL | 119 | 8.8 (7.0, 12.4) |
| Serum BD-tau (Simoa), pg/mL | 119 | 9.9 (7.5, 14.1) |
| Plasma GFAP (Simoa), pg/mL | 119 | 87 (57, 119) |
| Serum GFAP (Simoa), pg/mL | 119 | 103 (64, 149) |
| Data are presented as median (IQR) for continuous variables and n (%) for categorical variables. N indicates the number of participants included in each analysis. |  |  |

The correlations between biomarkers are shown in eFigure 1. Strong correlations were observed between NfL measurements obtained using different analytical platforms (Spearman’s *r* ≥ 0.93). Correlations between the different p-tau isoforms were moderate to strong (Spearman’s *r* = 0.56–0.90). In contrast, correlations between GFAP and the other blood-based biomarkers were generally weak (Spearman’s *r* = 0.07–0.33).

### Associations of blood-based biomarkers with demographic variables

Associations between blood-based biomarkers and demographic variables, including age, BMI and renal function, are summarized in eTable 3.

Spearman correlation analyses using non-transformed biomarker concentrations identified several significant associations with demographic variables. Older age was associated with higher serum p-tau217, plasma and serum BD-tau, and plasma and serum GFAP concentrations. Additionally, lower eGFR, indicating worse renal function, was associated with higher plasma and serum GFAP concentrations. Nominal associations that did not survive false discovery rate (FDR) correction were also observed between older age and higher plasma and serum NfL (Lumipulse), serum NfL (singleplex, Simoa) and serum NfL (N4PD, Simoa) concentrations; higher BMI with lower plasma and serum GFAP; and lower eGFR with higher serum BD-tau values.

Demographic characteristics stratified by sex are shown in eTable 4. In multivariable-adjusted models (Model 3), male patients showed higher plasma and serum p-tau181 and p-tau217, whereas female patients showed higher plasma GFAP (eTable 5). However, when analyzing the exploratory sensitivity model additionally adjusted for the number of LMN-affected regions (Model 4), only the associations with plasma and serum p-tau217 remained significant.

Demographic characteristics stratified by form of onset are shown in eTable 6. Bulbar-onset ALS was associated with higher plasma and serum NfL accross analytical platforms and higher plasma GFAP in unadjusted analyses (Model 1). In the multivariable-adjusted model (Model 3), bulbar-onset ALS was associated with lower plasma p-tau217. None of these associations remained significant after FDR correction (eTable 7, Model 3).

Demographic characteristics stratified by riluzole treatment status are shown in eTable 8. Riluzole-treated patients had longer survival (eTable 8), but no significant differences in blood biomarkers were observed between patients treated and not treated with riluzole (eTable 9, Models 1-3).

### Associations of blood-based biomarkers with disease severity

In cross-sectional analyses, higher p-tau181, p-tau217, BD-tau, as well as GFAP, in both plasma and serum, were associated with lower ALSFRS-R scores, indicating greater functional impairment (all q < 0.05, Figure 1 and eTable10, Model 3). Among tau biomarkers, p-tau217 and BD-tau showed the largest effect estimates for ALSFRS-R scores. Although plasma and serum NfL of different platforms were associated with ALSFRS-R in initial models (Models 1 and 2), these associations did not remain significant after adjustment for clinical covariates and after FDR correction (Figure 1, eTable 10, Model 3).

**Figure 1.**
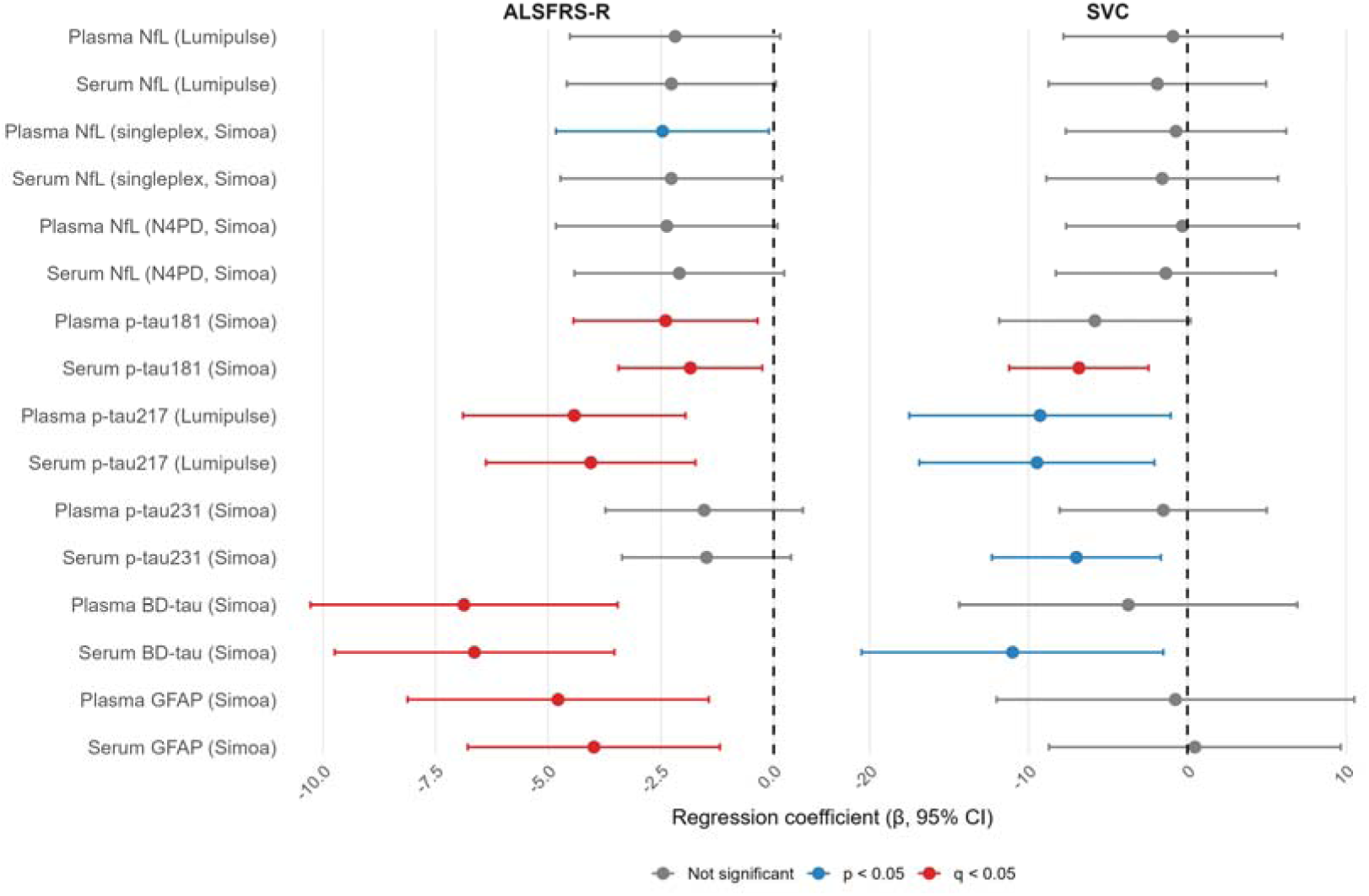
Associations between blood-based biomarkers and cross-sectional disease severity measures in ALS. Forest plots show multivariable-adjusted linear regression coefficients (β) and 95% confidence intervals for the associations between the log-transformed blood-based biomarkers and ALSFRS-R score (left panel) and SVC (right panel). Negative β coefficients indicate greater disease severity (i.e. lower ALSFRS-R or SVC) associated with higher biomarker values. Models were adjusted (according to Model 3) for age, sex, BMI, eGFR, ALS onset form, riluzole treatment and King’s stage (for ALSFRS-R analyses, left panel), and additionally for ALSFRS-R score (for SVC analyses, right panel). Blue symbols indicate nominal significance (p < 0.05), red symbols indicate significance after FDR correction (q < 0.05), and grey symbols indicate non-significant associations. Abbreviations: ALS = Amyotrophic Lateral Sclerosis; ALSFRS-R = Amyotrophic Lateral Sclerosis Functional Rating Scale–Revised; BD-tau = brain-derived tau; BMI = body mass index; eGFR = estimated glomerular filtration rate; FDR = false discovery rate; GFAP = glial fibrillary acidic protein; N4PD = Neurology 4-Plex D; NfL = neurofilament light chain; p-tau = phosphorylated tau; SVC = slow vital capacity.

Associations with SVC were more limited. Higher serum p-tau181 was associated with lower SVC (FDR q-value < 0.05, Figure 1 and eTable11, Model 3). Additionally, p-tau217, p-tau231 and BD-tau were associated with lower SVC in the same adjusted model; but did not remain significant after FDR correction (Figure 1, eTable 11, Model 3).

Overall, tau-related biomarkers, particularly p-tau217 and BD-tau, showed the strongest cross-sectional associations with disease severity. In contrast, associations between NfL and disease severity were attenuated after adjustment for clinical covariates and did not remain significant after FDR correction (Figure 1).

### Associations of blood-based biomarkers with lower motor neuron and muscle involvement

Higher plasma and serum p-tau181, p-tau217, p-tau231 and BD-tau were associated with higher CK and hs-cTnT, after multivariable adjustment and FDR correction (Figure 2, eTables 12 and 13, Model 3).

**Figure 2.**
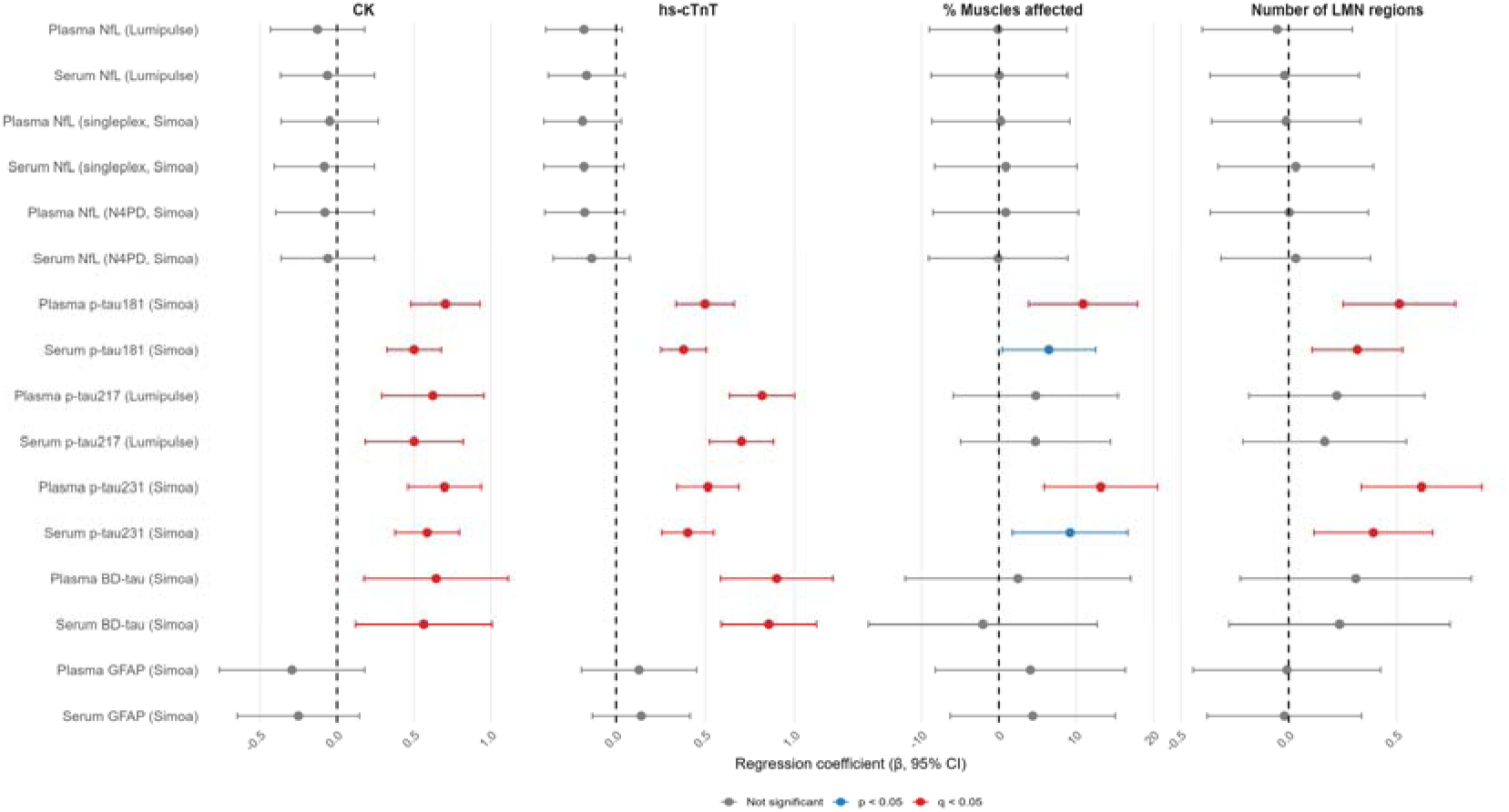
Associations between blood-based biomarkers and muscle/LMN involvement. Forest plots show multivariable-adjusted linear regression coefficients (β) and 95% confidence intervals for the associations between the log-transformed blood-based biomarkers and CK (first panel), hs-cTnT (second panel), percentage of affected muscles (third panel) and the number of LMN-affected regions (fourth panel). Positive β coefficients indicate greater muscle injury (i.e. higher CK or hs-cTnT), or a greater percentage of affected muscles and number of LMN-affected regions associated with higher biomarker concentrations. Models were adjusted (according to Model 3) for age, sex, BMI, eGFR, ALS onset form, riluzole treatment, King’s stage and ALSFRS-R score. Blue symbols indicate nominal significance (*p* < 0.05), red symbols indicate significance after false discovery rate (FDR) correction (q < 0.05), and grey symbols indicate non-significant associations. Analyses involving the proportion of affected muscles and the number of LMN-affected regions were restricted to participants with available EMG data (n = 67). Abbreviations: ALS = Amyotrophic Lateral Sclerosis; ALSFRS-R = Amyotrophic Lateral Sclerosis Functional Rating Scale–Revised; BD-tau = brain-derived tau; BMI = body mass index; CK = creatine kinase; eGFR = estimated glomerular filtration rate; FDR = false discovery rate; GFAP = glial fibrillary acidic protein; hs-cTnT = high-sensitivity cardiac troponin T; LMN = lower motor neuron; N4PD = Neurology 4-Plex D; NfL = neurofilament light chain; p-tau = phosphorylated tau.

Higher plasma and serum p-tau181 and p-tau231 were also associated with greater LMN involvement, measured as the number of LMN-affected regions (eTable 14, Model 3) and the proportion of affected muscles (eTable 15, Model 3). These associations remained significant after multivariable adjustment and FDR correction (q < 0.05), except for the associations of serum p-tau181 and p-tau231 with the proportion of affected muscles. Associations for plasma and serum p-tau217 and BD-tau with LMN involvement were also observed, but they did not remain significant after further adjustment and FDR correction (Figure 2, eTables 14 and 15, Model 2). In contrast, plasma and serum NfL, across all analytical platforms, and GFAP were not associated with CK, hs-cTnT or LMN involvement (Models 1-3).

Overall, these findings indicate a closer association of tau-related biomarkers with LMN and muscle-related disease burden in ALS.

### Associations of blood-based biomarkers with disease progression and survival

Higher plasma and serum NfL, across all analytical platforms, were consistently associated with faster disease progression, measured by ALSFRS-R slope, after multivariable adjustment and FDR correction (all FDR-corrected q < 0.001, Model 3). Although higher plasma and serum BD-tau were also associated with faster disease progression (Models 1-2), they did not remain significant after multivariable adjustment and FDR correction (Model 3). No significant associations were observed for the remaining tau-related biomarkers or GFAP (Figure 3, eTable 16, Model 3).

**Figure 3.**
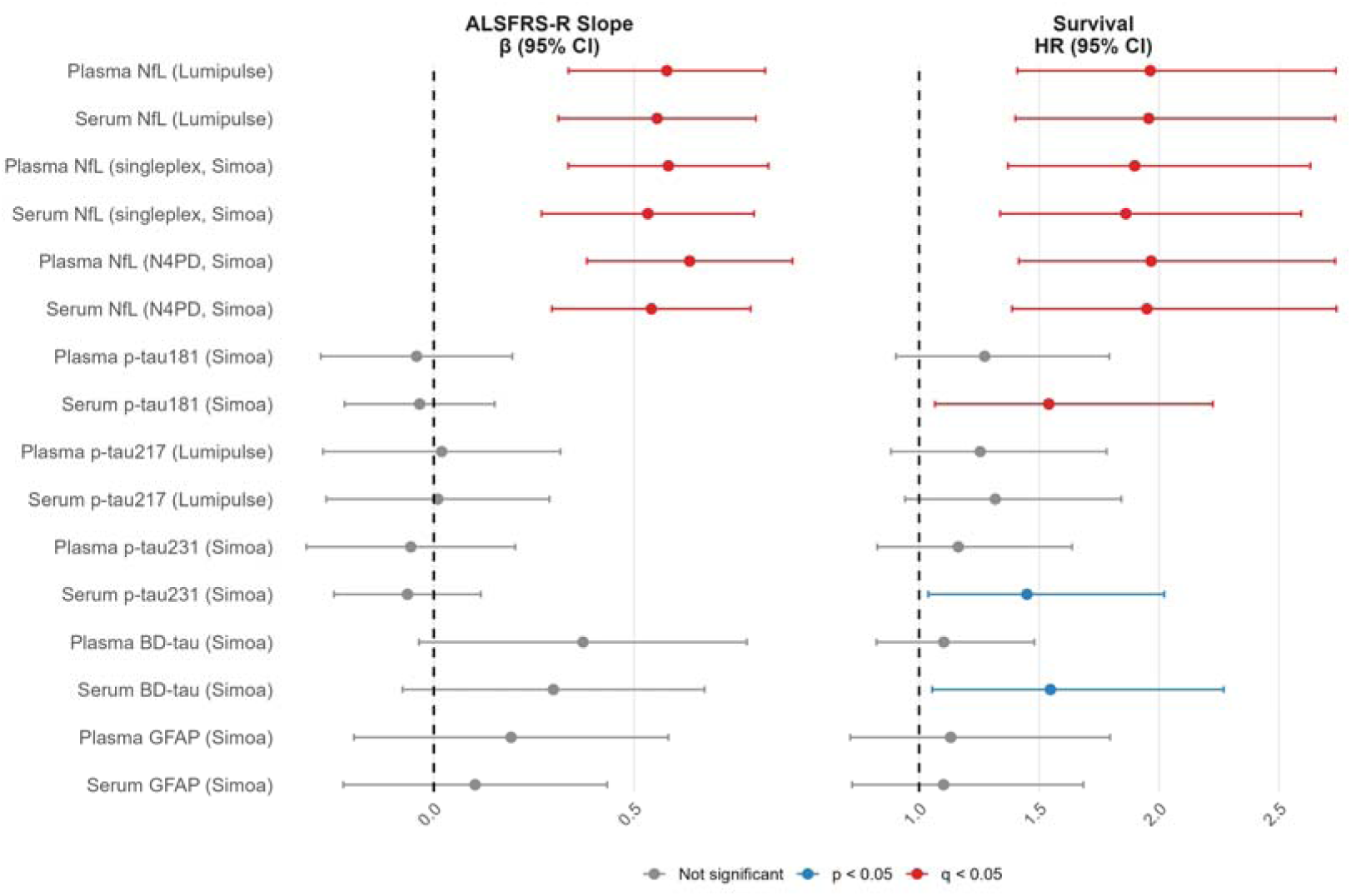
Associations between blood-based biomarkers and disease progression and survival in ALS. Forest plots show multivariable-adjusted linear regression coefficients (β) and 95% confidence intervals for associations between the log-transformed blood-based biomarkers with ALSFRS-R slope (left panel), and hazard ratios (HRs) with 95% confidence intervals for associations of the log-transformed and standardized (z-score) blood-based biomarkers with survival (right panel). Positive β coefficients indicate faster disease progression. HRs >1 indicate decreased risk of survival with higher biomarker values. Models were adjusted (according to Model 3) for age, sex, BMI, eGFR, ALS onset form, riluzole treatment, King’s stage and baseline ALSFRS-R score. Blue symbols indicate nominal significance (p < 0.05), red symbols indicate significance after false discovery rate (FDR) correction (q < 0.05), and grey symbols indicate non-significant associations. Abbreviations: ALS = Amyotrophic Lateral Sclerosis; ALSFRS-R = Amyotrophic Lateral Sclerosis Functional Rating Scale–Revised; BD-tau = brain-derived tau; BMI = body mass index; eGFR = estimated glomerular filtration rate; FDR = false discovery rate; GFAP = glial fibrillary acidic protein; HR = hazard ratio; N4PD = Neurology 4-Plex D; NfL = neurofilament light chain; p-tau = phosphorylated tau.

Higher plasma and serum NfL were also consistently associated with reduced survival across all models (all q < 0.001, Models 1-3). Nominal associations with survival were observed for serum p-tau181, p-tau231 and BD-tau in adjusted models (Model 3); however, none remained significant after FDR correction (Figure 3, eTable 17, Model 3).

An exploratory sensitivity analysis was additionally adjusted for the extent of LMN involvement (Model 4). In this multivariable-adjusted model, plasma and serum NfL, as well as serum BD-tau, did not remain significant after FDR correction, although q-values remained close to the significance threshold for NfL (q = 0.053; eTable 17, Model 4).

Overall, NfL was the only biomarker independently associated with both disease progression and survival, whereas tau-related biomarkers were not independently associated with longitudinal outcomes after adjustment.

## Discussion

In this longitudinal observational study of 119 well characterized patients with ALS, blood-based tau biomarkers and NfL showed distinct clinical associations. Tau-related biomarkers, particularly p-tau217 and BD-tau, were consistently associated with cross-sectional disease severity, whereas p-tau181 and p-tau231 were most closely related to LMN and muscle involvement. In contrast, NfL was the only biomarker independently associated with both disease progression and survival, confirming its role as a robust prognostic marker in ALS. Together, these findings indicate that different blood-based biomarkers capture distinct biological and clinical aspects of ALS: tau-related biomarkers preferentially reflect cross-sectional severity and LMN and muscle-related disease burden, whereas NfL captures disease aggressiveness and prognosis.

While previous studies have largely focused on whether blood tau biomarkers are altered in ALS, our study addressed a different question: which clinically relevant characteristics of ALS are reflected by different blood-based biomarkers. By simultaneously assessing multiple tau-related biomarkers in plasma and serum, alongside NfL and GFAP, and relating them to quantitative measures of LMN involvement, muscle injury, functional status, disease progression and survival, we were able to delineate distinct biomarker profiles associated with different aspects of disease expression. These analyses revealed that individual tau-related biomarkers show distinct clinical associations that differ from those of NfL.

Previous work showed that plasma p-tau181 is increased in ALS independently of Alzheimer’s disease pathology and may relate to LMN involvement. Vacchiano *et al*. reported elevated plasma p-tau181 in ALS despite no corresponding CSF elevation and no clear relationship with AD biomarkers.^26^ More recently, Abu-Rumeileh *et al*. showed that serum p-tau181 and p-tau217 are increased in ALS and that both species are detectable in skeletal muscle, with increased immunoreactivity in atrophic muscle fibers.^29^

The association between tau-related biomarkers and LMN or muscle-related measures supports a possible peripheral contribution to blood tau elevations in ALS. This interpretation is consistent with the absence of parallel CSF p-tau increases in prior studies and with evidence of p-tau181 and p-tau217 expression in atrophic ALS muscle fibers.^29,51^ The strongest LMN associations in our cohort were observed for p-tau181 and p-tau231, whereas p-tau217 and BD-tau were more closely associated with global functional impairment.^15,30^ This suggests that individual tau species may not be interchangeable in ALS and may capture partially different aspects of disease expression.

BD-tau has been proposed as a blood biomarker enriched for brain-derived tau species and developed to avoid peripheral “big tau” isoforms.^41^ In AD cohorts, BD-tau has been associated with amyloid-linked neurodegeneration and cognitive decline.^52^ In ALS, however, the clinical relevance of BD-tau remains less established.^28^ Our finding that BD-tau was associated with ALSFRS-R but not robustly with LMN involvement suggests that BD-tau may capture disease severity through mechanisms that may differ from those reflected by phosphorylated tau species.

Thus, our findings provide additional evidence supporting the utility of blood p-tau biomarkers as markers of LMN involvement in ALS, while identifying BD-tau as a complementary marker of disease severity alongside NfL. Further studies integrating paired CSF, blood, muscle, neurophysiological and neuropathological data will be required to clarify the biological origin and pathophysiological significance of the different circulating tau species in ALS.

In contrast to tau-related biomarkers, NfL was the only biomarker consistently associated with disease progression and survival. This is consistent with prior literature supporting NfL as a marker of axonal injury and a robust prognostic biomarker in ALS.^4,12,15,19,53^ In our study, NfL was associated with faster ALSFRS-R decline and shorter survival time but was not independently associated with LMN or muscle involvement. These findings suggest that NfL mainly reflects overall disease aggressiveness rather than the anatomical distribution of motor neuron involvement.

GFAP showed more limited associations. Although GFAP was associated with ALSFRS-R, it was not associated with CK, hs-cTnT, LMN involvement, disease progression or survival. Thus, GFAP did not provide the same phenotype-specific information as tau-related biomarkers or the prognostic information provided by NfL. Recent work has evaluated GFAP together with NfL and p-tau181 in ALS, but its independent clinical utility remains less clear.^33,34^ In contrast, plasma GFAP may indicate the presence of concomitant AD neuropathological changes in a subset of patients with ALS ^54,55^

Sex- and onset-related differences in biomarkers were largely attenuated after multivariable adjustment, indicating that these associations were not independent of other demographic and clinical characteristics.

Overall, our results provide new evidence regarding the potential origin and utility of blood-related tau biomarkers in ALS and their complementary role to NfL. While NfL represents a robust diagnostic and prognostic biomarker, tau-related biomarkers may provide additional information for patient phenotyping according to the degree of peripheral impairment and clinical status. The combined assessment of tau-related biomarkers and NfL may improve biological and clinical characterization of ALS and could help inform patient stratification in future observational studies and clinical trials. Our findings further support the concept that ALS biomarker biology is multidimensional, with different biomarker classes reflecting partially distinct biological processes.

Limitations include the single-center design, missing SVC, EMG and CK data in a subset of participants, and the absence of paired CSF, which precluded determination of CSF-based AT(N) status, neuropsychological assessment, neuroimaging, muscle biopsy or neuropathologic measures. We also did not quantify upper motor neuron burden, limiting assessment of whether specific biomarkers differentially capture UMN vs LMN involvement. Finally, EMG study and blood draw were not conducted simultaneously, and EMG-derived measures of LMN involvement were available only in a subset of participants. Several strengths should be noted. The cohort was clinically well characterized, included survival follow-up, and allowed comparison of multiple blood biomarkers across plasma and serum using different analytical platforms. In addition, the analyses examined several clinically relevant ALS features, including functional status, respiratory function, progression rate, survival, LMN involvement and muscle injury markers.

In conclusion, blood-based biomarkers in ALS capture different aspects of the disease. Tau-related biomarkers were associated with cross-sectional disease severity, LMN involvement and muscle injury, whereas NfL was independently associated with disease progression and survival. These findings support the complementary use of tau-related biomarkers and NfL for ALS phenotypic characterization and prognosis assessement, respectively.

## Acknowledgements

The authors sincerely thank the ELAMAR participants and their families, as well as the Catalan ALS Foundation (Fundació Miquel Valls), without whom this research would not have been possible. We also thank the staff of the Neurology Department at Hospital del Mar and the Barcelonaβeta Brain Research Center for their technical and logistical support; and Marc Sebastián Romagosa for the statistical review. In addition, we thank Fujirebio and Quanterix for providing the kits used for plasma and serum biomarker measurements, and ADx (Eugeen Vanmechelen) for providing the ADx253 antibody.

During the preparation of this work, the authors used AI tools to summarize the text and review the grammar. After using this tool, the authors reviewed and edited the content as needed and take full responsibility for the content of the publication.

## Glossary

AD: Alzheimer’s disease
ALS: amyotrophic lateral sclerosis
ALSFRS-R: Amyotrophic Lateral Sclerosis Functional Rating Scale–Revised
ANCOVA: analysis of covariance
ASO: antisense oligonucleotide
BBRC: Barcelonaβeta Brain Research Center
BD-tau: brain-derived tau
BMI: body mass index
CI: confidence interval
CK: creatine kinase
CSF: cerebrospinal fluid
CV: coefficient of variation
eGFR: estimated glomerular filtration rate
EMG: electromyography
FDR: false discovery rate
FTD: frontotemporal dementia
GFAP: glial fibrillary acidic protein
HR: hazard ratio
hs-cTnT: high-sensitivity cardiac troponin
T iQC: internal quality control
IQR: interquartile range
LMN: lower motor neuron
N4PD: Neurology 4-Plex
D NfL: neurofilament light chain
PH: proportional hazard
p-tau: phosphorylated tau
SVC: slow vital capacity
UMN: upper motor neuron

## Author Contributors

BB-R: drafting/revision of the manuscript for content; major role in the acquisition of data; study concept or design; analysis or interpretation of the data. PO-R: drafting/revision of the manuscript for content; major role in the acquisition of data; technical assistance in sample processing; analysis or interpretation of the data. FL-H: drafting/revision of the manuscript for content; major role in the acquisition of data; analysis or interpretation of the data; database management. SVN: revision of the manuscript for content; major role in the acquisition of data. MD-O: revision of the manuscript for content, technical assistance in sample processing.; HB-F: revision of the manuscript for content, technical assistance in sample processing. EJ-M: revision of the manuscript for content, technical assistance in sample processing. MLT: revision of the manuscript for content; JT-T: revision of the manuscript for content. MC: revision of the manuscript for content. MaRP: revision of the manuscript for content; major role in the acquisition of data; study concept or design; analysis or interpretation of the data. MS-C: drafting/revision of the manuscript for content, including medical writing for content; study concept or design; analysis or interpretation of the data.

## Supplemental Material

**eFigure 1.**
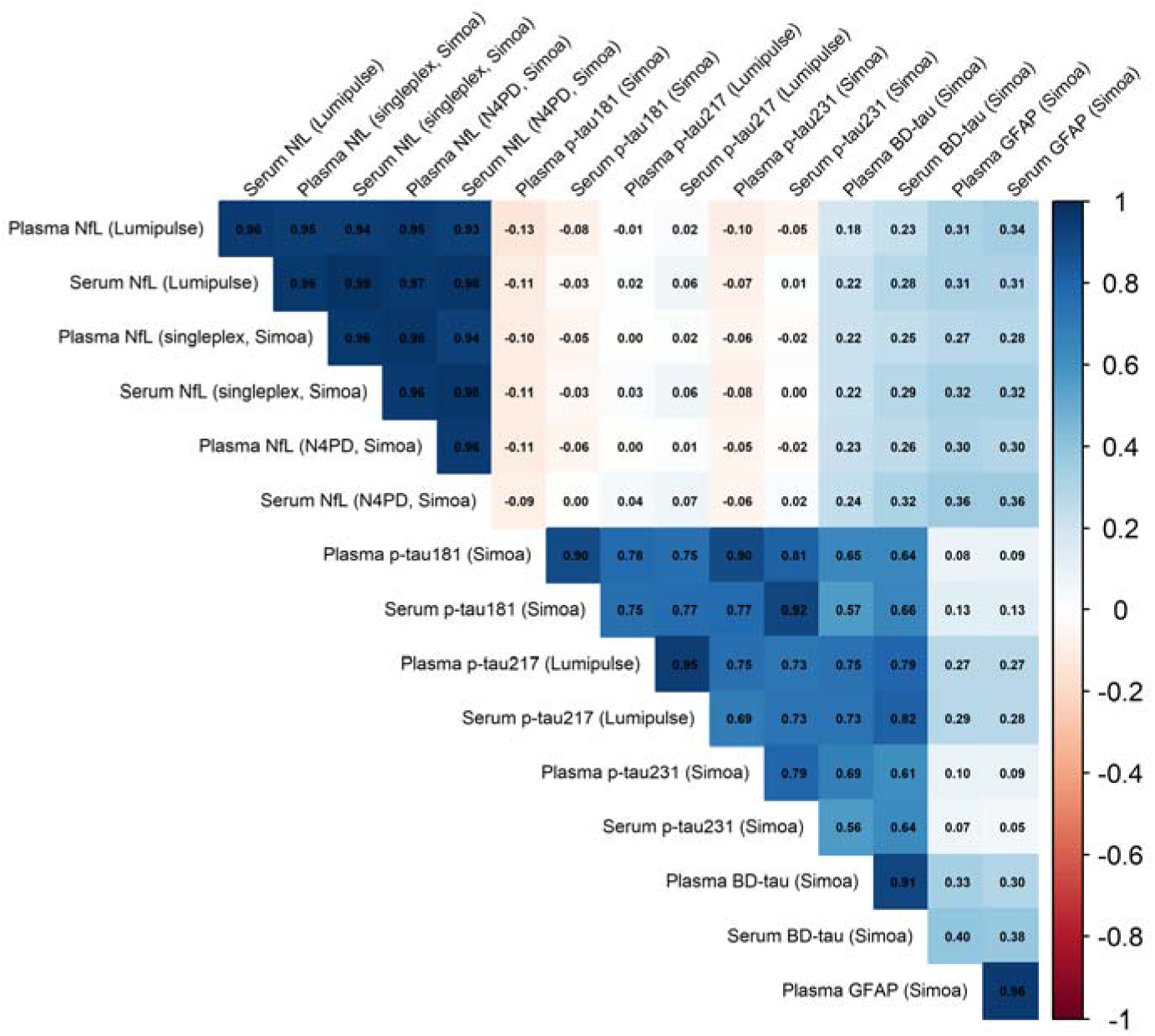
Correlation matrix between blood-based biomarkers. Spearman’s rank correlation coefficients are shown. Cell color represents the magnitude and direction of the Spearman correlation coefficient, with darker blue indicating stronger positive correlations, darker red indicating stronger negative correlations; and lighter colors indicating weaker correlations. Correlations were considered statistically significant at *p* < 0.05. All pairwise correlations were statistically significant except those between plasma and serum NfL (all technologies) with plasma and serum p-tau181, p-tau217 and p-tau231; and between plasma and serum GFAP with plasma and serum p-tau181 and p-tau231. Abbreviations: BD-tau = brain-derived tau; GFAP = glial fibrillary acidic protein; N4PD = Neurology 4-Plex D; NfL = neurofilament light chain; p-tau = phosphorylated tau.

**eTable 1:** Inter-run and intra-run %CV.

| Assay | N | Inter-run %CV | Intra-run %CV |
| --- | --- | --- | --- |
| Plasma NfL (Lumipulse) | 119 | 9.09 | 12.87 |
| Serum NfL (Lumipulse) |  |  |  |
| Plasma NfL (singleplex, Simoa) | 119 | 15.43 | 11.78 |
| Serum NfL (singleplex, Simoa) | 119 | 7.25 | 12.58 |
| Plasma NfL (N4PD, Simoa) | 119 | 15.70 | 11.85 |
| Serum NfL (N4PD, Simoa) | 119 | 17.46 | 5.72 |
| Plasma p-tau181 (Simoa) | 118 | 7.14 | 8.23 |
| Serum p-tau181 (Simoa) | 116 | 7.08 | 7.87 |
| Plasma p-tau217 (Lumipulse) | 119 | 12.16 | 8.29 |
| Serum p-tau217 (Lumipulse) |  |  |  |
| Plasma p-tau231 (Simoa) | 116 | 10.29 | 6.40 |
| Serum p-tau231 (Simoa) | 114 | 6.25 | 4.55 |
| Plasma BD-tau (Simoa) | 119 | 7.57 | 6.23 |
| Serum BD-tau (Simoa) | 119 | 10.35 | 6.02 |
| Plasma GFAP (Simoa) | 119 | 11.75 | 7.00 |
| Serum GFAP (Simoa) | 119 | 8.09 | 5.50 |
| Inter- and intra-run coefficients of variation (%CV) for each biomarker are presented. N indicates the number of participants included in each analysis.<br>Abbreviations: BD-tau = brain-derived tau; GFAP = glial fibrillary acidic protein; N4PD = Neurology 4-Plex D; NfL = neurofilament light chain; p-tau = phosphorylated tau. |  |  |  |

**eTable 2:** STROBE statement completed checklist.

|  | Item Description | Location (or reason for not reporting) |
| --- | --- | --- |
| **Title and abstract** |  |  |
| 1a. Indicate the study's design | Indicate the study's design with a commonly used term in the title or the abstract. | Abstract, Methods |
| 1b. Abstract | Provide in the abstract an informative and balanced summary of what was done and what was found. | Abstract, Methods and Results |
| **Introduction** |  |  |
| 2. Background / rationale | Explain the scientific background and rationale for the investigation being reported. | Introduction; Paragraph 1-5 |
| 3. Objectives | State specific objectives, including any prespecified hypotheses. | Introduction; Paragraph 6 |
| **Methods** |  |  |
| 4. Study design | Present key elements of study design early in the paper. | Methods; Study design and participants |
| 5. Setting | Describe the setting, locations, and relevant dates, including periods of recruitment, exposure, follow-up, and data collection. | Methods; Study design and participants |
| 6a. Eligibility criteria | **Cohort study:** Give the eligibility criteria, and the sources and methods of selection of participants. Describe methods of follow-up. **Case-control study:** Give the eligibility criteria, and the sources and methods of case ascertainment and control selection. Give the rationale for the choice of cases and controls. **Cross-sectional study:** Give the eligibility criteria, and the sources and methods of selection of participants. | Methods; Study design and participants |
| 6b. Matching criteria | **Cohort study:** For matched studies, give matching criteria and number of exposed and unexposed. **Case-control study:** For matched studies, give matching criteria and the number of controls per case. | - |
| 7. Variables | Clearly define all outcomes, exposures, predictors, potential confounders, and effect modifiers. Give diagnostic criteria, if applicable. | Methods; Clinical assessments and, Sample processing and biomarker measurement |
| 8. Data sources / measurement | For each variable of interest give sources of data and details of methods of assessment (measurement). Describe comparability of assessment methods if there is more than one group. | Methods; Clinical assessments and, Sample processing and biomarker measurement |
| 9. Bias | Describe any efforts to address potential sources of bias. | Methods; Clinical assessments and, Sample processing and biomarker measurement |
| 10. Study size | Explain how the study size arrived. | Methods; Study design and participants |
| 11. Quantitative variables | Explain how quantitative variables were handled in the analyses. If applicable, describe which groupings were chosen, and why. | Methods; Clinical assessments and, Sample processing and biomarker measurement |
| 12a. Statistical methods | Describe all statistical methods, including those used to control for confounding. | Methods; Statistical analyses |
| 12b. Statistical methods – subgroups and interactions | Describe any methods used to examine subgroups and interactions. | Methods; Statistical analyses |
| 12c. Statistical methods – missing data | Explain how missing data were addressed. | Methods; Statistical analyses |
| 12di. Statistical methods – loss to follow-up | **Cohort study:** If applicable, describe how loss to follow-up was addressed. | Methods; Statistical analyses; Paragraph 6 |
| 12dii. Statistical methods – matching cases and controls | **Case-control study:** If applicable, explain how matching of cases and controls was addressed. | - |
| 12diii. Statistical methods – sampling strategy | **Cross-sectional study:** If applicable, describe analytical methods taking account of sampling strategy. | - |
| 12e. Statistical methods – sensitivity analyses | Describe any sensitivity analyses. | Methods; Statistical analyses |
| **Results** |  |  |
| 13a. Participant numbers | Report the numbers of individuals at each stage of the study—e.g., numbers potentially eligible, examined for eligibility, confirmed eligible, included in the study, completing follow-up, and analysed; Consider use of a flow diagram. | Methods; Study design and participants |
| 13b. Participants – non-participation | Give reasons for non-participation at each stage. | - |
| 13c. Participants – flow diagram | Consider use of a flow diagram. | - |
| 14a. Descriptive data – participant characteristics | Give characteristics of study participants (e.g., demographic, clinical, social) and information on exposures and potential confounders. Present the information in a table. | Results; Demographics; Table 1; eTable 3 |
| 14b. Descriptive data – missing data | Indicate the number of participants with missing data for each variable of interest. | Results; Demographics; Table 1 |
| 14c. Descriptive data – follow-up time | **Cohort study:** Summarise follow-up time—e.g., average and total amount. | Results; Demographics; Table 1 |
| 15. Outcome data | **Cohort study:** Report numbers of outcome events or summary measures over time. **Case-control study:** Report numbers in each exposure category, or summary measures of exposure. **Cross-sectional study:** Report numbers of outcome events or summary measures. | Results; Demographics; Table 1 |
| 16a. Main results | Give unadjusted estimates and, if applicable, confounder-adjusted estimates and their precision (e.g., 95% confidence intervals). Make clear which confounders were adjusted for and why they were included. | Methods; Statistical analyses; Paragraph 4<br>Results; all. |
| 16b. Main results – category boundaries | Report category boundaries when continuous variables were categorised. | - |
| 16c. Main results – risk | If relevant, consider translating estimates of relative risk into absolute risk for a meaningful time period. | - |
| 17. Other analyses | Report other analyses done—e.g., analyses of subgroups and interactions, and sensitivity analyses. | Methods; Statistical analyses<br>Results; all |
| **Discussion** |  |  |
| 18. Key results | Summarise key results with reference to study objectives. | Discussion; Paragraph 1 and 2 |
| 19. Limitations | Discuss limitations of the study, taking into account sources of potential bias or imprecision. Discuss both direction and magnitude of any potential bias. | Discussion; Paragraph 11 |
| 20. Interpretation | Give a cautious overall interpretation considering objectives, limitations, multiplicity of analyses, results from similar studies, and other relevant evidence. | Discussion; Paragraph 10 |
| 21. Generalisability | Discuss the generalisability (external validity) of the study results. | - |
| **Other information** |  |  |
| 22. Funding | Give the source of funding and the role of the funders for the present study and, if applicable, for the original study on which the present article is based. | Study Funding |

**eTable 3:** Correlations between biomarkers and clinical covariates.

| Biomarker | Age |  |  |  | BMI |  |  |  | eGFR |  |  |  |
| --- | --- | --- | --- | --- | --- | --- | --- | --- | --- | --- | --- | --- |
|  | N | r | p-value | FDR q-value | N | r | p-value | FDR q-value | N | r | p-value | FDR q-value |
| Plasma NfL (Lumipulse) | 119 | 0.22 | **0.016** | 0.075 | 119 | -0.12 | 0.195 | 0.323 | 117 | -0.13 | 0.170 | 0.291 |
| Serum NfL (Lumipulse) | 119 | 0.21 | **0.025** | 0.085 | 119 | -0.13 | 0.147 | 0.272 | 117 | -0.11 | 0.219 | 0.350 |
| Plasma NfL (singleplex, Simoa) | 119 | 0.16 | 0.074 | 0.187 | 119 | -0.17 | 0.066 | 0.187 | 117 | -0.10 | 0.276 | 0.390 |
| Serum NfL (singleplex, Simoa) | 119 | 0.22 | **0.019** | 0.075 | 119 | -0.15 | 0.107 | 0.234 | 117 | -0.14 | 0.139 | 0.272 |
| Plasma NfL (N4PD, Simoa) | 119 | 0.17 | 0.071 | 0.187 | 119 | -0.15 | 0.107 | 0.234 | 117 | -0.10 | 0.276 | 0.390 |
| Serum NfL (N4PD, Simoa) | 119 | 0.22 | **0.016** | 0.075 | 119 | -0.14 | 0.126 | 0.263 | 117 | -0.13 | 0.165 | 0.291 |
| Plasma p-tau181 (Simoa) | 118 | 0.14 | 0.145 | 0.272 | 118 | 0.08 | 0.399 | 0.504 | 116 | 0.00 | 0.969 | 0.983 |
| Serum p-tau181 (Simoa) | 116 | 0.16 | 0.093 | 0.223 | 116 | 0.10 | 0.296 | 0.406 | 114 | -0.04 | 0.636 | 0.726 |
| Plasma p-tau217 (Lumipulse) | 119 | 0.22 | **0.018** | 0.075 | 119 | 0.00 | 0.979 | 0.983 | 117 | -0.04 | 0.701 | 0.747 |
| Serum p-tau217 (Lumipulse) | 119 | 0.24 | **0.008** | **0.052** | 119 | 0.00 | 0.983 | 0.983 | 117 | -0.09 | 0.346 | 0.449 |
| Plasma p-tau231 (Simoa) | 116 | 0.09 | 0.336 | 0.448 | 116 | 0.06 | 0.538 | 0.630 | 114 | 0.04 | 0.693 | 0.747 |
| Serum p-tau231 (Simoa) | 114 | 0.07 | 0.469 | 0.563 | 114 | 0.11 | 0.264 | 0.390 | 112 | 0.04 | 0.685 | 0.747 |
| Plasma BD-tau (Simoa) | 119 | 0.29 | **0.001** | **0.010** | 119 | 0.10 | 0.275 | 0.390 | 117 | -0.17 | 0.070 | 0.187 |
| Serum BD-tau (Simoa) | 119 | 0.32 | **<0.001** | **0.003** | 119 | 0.08 | 0.411 | 0.505 | 117 | -0.20 | **0.033** | 0.104 |
| Plasma GFAP (Simoa) | 119 | 0.67 | **<0.001** | **<0.001** | 119 | -0.21 | **0.022** | 0.081 | 117 | -0.48 | **<0.001** | **<0.001** |
| Serum GFAP (Simoa) | 119 | 0.63 | **<0.001** | **<0.001** | 119 | -0.22 | **0.017** | 0.075 | 117 | -0.44 | **<0.001** | **<0.001** |

Spearman rank correlation coefficients (*r*) and corresponding two-sided p-values, as well as the FDR corrected q-values, are presented, with p-values and q-values < 0.05 shown in bold. N indicates the number of participants included in each analysis. Correlations were calculated using non-transformed biomarker concentrations. Abbreviations: BD-tau = brain-derived tau; BMI = body mass index; eGFR = estimated glomerular filtration rate; GFAP = glial fibrillary acidic protein; N4PD = Neurology 4-Plex D; NfL = neurofilament light chain; p-tau = phosphorylated tau.

**eTable 4:** Demographics by sex.

| Characteristic | N | Female<br>(N=67) | Male<br>(N=52) | p-value |
| --- | --- | --- | --- | --- |
| Age, years | 119 | 66 (57, 74) | 63 (52, 74) | 0.450 |
| BMI, kg/m<sup>2</sup> | 119 | 22.67 (18.60, 25.10) | 24.35 (21.53, 27.33) | 0.052 |
| eGFR, mL/min/1.73 m<sup>2</sup> | 117 | 103.15 (92.40, 111.35) | 105.71 (94.06, 119.82) | 0.112 |
| ALS onset form | 119 |  |  | 0.074 |
| Bulbar |  | 27 (40%) | 12 (23%) |  |
| Spinal |  | 40 (60%) | 40 (77%) |  |
| King's stage | 119 |  |  | **0.047** |
| Kings 1/2/3 |  | 24 (36%) | 29 (56%) |  |
| Kings 4 |  | 43 (64%) | 23 (44%) |  |
| ALSFRS-R, points | 119 | 31 (23, 40) | 34.50 (26.50, 40) | **0.040** |
| ALSFRS-R slope, points lost/month | 119 | 0.90 (0.56, 1.42) | 0.82 (0.44, 1.61) | 0.789 |
| Riluzole treatment | 119 | 47 (70%) | 26 (50%) | **0.040** |
| Months treated | 73 | 6 (3, 12) | 6 (3, 12) | 0.885 |
| Survival time from symptom onset, months | 119 | 32.42 (20.93, 47.48) | 29.81 (19.49, 42.97) | 0.402 |
| Death during follow-up | 119 | 42 (63%) | 29 (56%) | 0.566 |
| SVC, % predicted | 81 | 70 (51, 81) | 77.85 (62, 90) | 0.110 |
| CK, U/L | 90 | 82 (57, 136) | 194.50 (98, 370) | **<0.001** |
| hs-cTnT, ng/L | 113 | 16.67 (9.46, 28.65) | 27.67 (18.97, 50.65) | **<0.001** |
| Data are presented as median (IQR) for continuous variables and n (%) for categorical variables. N indicates the number of participants included in each analysis. P-values for continuous variables were calculated using the Wilcoxon rank-sum test, and p-values for categorical variables were calculated using Pearson's chi-square test. Statistically significant p-values (p < 0.05) are shown in bold.<br>Abbreviations: ALS = amyotrophic lateral sclerosis; ALSFRS-R = Amyotrophic Lateral Sclerosis Functional Rating Scale-Revised; BMI = body mass index; CK = creatine kinase; eGFR = estimated glomerular filtration rate; hs-cTnT = high-sensitivity cardiac troponin T; SVC = Slow Vital Capacity. |  |  |  |  |

**eTable 5.** Associations between sex and blood-based biomarkers.

| Biomarker | Femal<br>(N=67) | Male<br>(N=52) | Model 1 |  | Model 2 |  | Model 3 |  |  | Model 4 |  |  |
| --- | --- | --- | --- | --- | --- | --- | --- | --- | --- | --- | --- | --- |
|  |  |  | N | p-value | N | p-value | N | p-value | FDR<br>q-value | N | p-value | FDR<br>q-value |
| Plasma NfL (Lumipulse) | 112.16<br>(69.15,<br>157.70) | 86.67<br>(51.30,<br>127.08) | 119 | **0.040** | 117 | 0.098 | 117 | 0.306 | 0.441 | 67 | 0.287 | 0.471 |
| Serum NfL (Lumipulse) | 117.71<br>(73.03,<br>157.04) | 82.52<br>(49.78,<br>135.16) | 119 | **0.030** | 117 | 0.091 | 117 | 0.331 | 0.441 | 67 | 0.318 | 0.471 |
| Plasma NfL (singleplex, Simoa) | 75.80<br>(44.00,<br>95.15) | 52.65<br>(33.22,<br>82.12) | 119 | 0.057 | 117 | 0.169 | 117 | 0.513 | 0.513 | 67 | 0.353 | 0.471 |
| Serum NfL (singleplex, Simoa) | 96.30<br>(61.45,<br>128.50) | 73.15<br>(42.23,<br>109.5) | 119 | **0.037** | 117 | 0.128 | 117 | 0.424 | 0.487 | 67 | 0.502 | 0.618 |
| Plasma NfL (N4PD, Simoa) | 67.76<br>(47.04,<br>92.66) | 54.62<br>(33.26,<br>77.64) | 119 | **0.045** | 117 | 0.136 | 117 | 0.426 | 0.487 | 67 | 0.348 | 0.471 |
| Serum NfL (N4PD, Simoa) | 99.67<br>(59.57,<br>127.58) | 74.6<br>(43.38,<br>112.4) | 119 | 0.06 | 117 | 0.16 | 117 | 0.482 | 0.513 | 67 | 0.636 | 0.678 |
| Plasma p-tau181 (Simoa) | 35.44<br>(22.89,<br>59.46) | 62.75<br>(29.72,<br>103.38) | 118 | **<0.001** | 116 | **0.002** | 116 | **<0.001** | **0.006** | 67 | **0.037** | 0.147 |
| Serum p-tau181 (Simoa) | 23.80<br>(11.90,<br>42.30) | 44.50<br>(25.60,<br>68.30) | 116 | **0.002** | 114 | **0.002** | 114 | **<0.001** | **0.006** | 65 | **0.028** | 0.147 |
| Plasma p-tau217 (Lumipulse) | 0.18<br>(0.12,<br>0.3) | 0.24<br>(0.16,<br>0.38) | 119 | **0.016** | 117 | **0.015** | 117 | **0.002** | **0.008** | 67 | **0.002** | **0.018** |
| Serum p-tau217 (Lumipulse) | 0.18<br>(0.13,<br>0.32) | 0.27<br>(0.16,<br>0.42) | 119 | **0.017** | 117 | **0.010** | 117 | **0.001** | **0.006** | 67 | **0.002** | **0.018** |
| Plasma p-tau231 (Simoa) | 3.78<br>(2.58,<br>6.03) | 5.21<br>(3.29,<br>8.00) | 116 | 0.115 | 116 | 0.283 | 116 | 0.197 | 0.315 | 66 | 0.598 | 0.673 |
| Serum p-tau231 (Simoa) | 1.53<br>(0.81,<br>2.66) | 2.34<br>(1.35,<br>3.7) | 114 | **0.009** | 114 | **0.030** | 114 | **0.032** | 0.086 | 64 | 0.173 | 0.346 |
| Plasma BD-tau (Simoa) | 8.76<br>(6.90,<br>11.10) | 9.10<br>(7.21,<br>13.40) | 119 | 0.333 | 117 | 0.433 | 117 | 0.053 | 0.105 | 67 | 0.107 | 0.286 |
| Serum BD-tau (Simoa) | 9.77<br>(7.18,<br>13.58) | 10.13<br>(7.67,<br>14.83) | 119 | 0.361 | 117 | 0.385 | 117 | 0.051 | 0.105 | 67 | 0.061 | 0.195 |
| Plasma GFAP (Simoa) | 97.88<br>(68.63,<br>129.31) | 65.54<br>(45.71,<br>99.84) | 119 | **<0.001** | 117 | **<0.001** | 117 | **0.007** | **0.022** | 67 | 0.160 | 0.346 |
| Serum GFAP (Simoa) | 115.3<br>(80.73,<br>156.32) | 73.93<br>(52.40,<br>119.78) | 119 | **0.003** | 117 | **0.009** | 117 | 0.059 | 0.105 | 67 | 0.767 | 0.767 |

Data are presented as median (IQR) in pg/mL. N indicates the number of participants included in each analysis. Group comparisons were performed using analysis of covariance (ANCOVA). Model 1: unadjusted. Model 2: adjusted for age, BMI and eGFR. Model 3: adjusted for age, BMI, eGFR, ALS onset form, riluzole treatment, King's stage and ALSFRS-R. Model 4: adjusted for age, BMI, eGFR, ALS onset form, riluzole treatment, King's stage, ALSFRS-R and number of LMN-affected regions. False discovery rate (FDR) q-values were calculated using the Benjamini-Hochberg procedure for Models 3 and 4. Statistically significant p- and q-values (<0.05) are shown in bold.
Abbreviations: ALS = amyotrophic lateral sclerosis; ALSFRS-R = Amyotrophic Lateral Sclerosis Functional Rating Scale-Revised; BD-tau = brain-derived tau; BMI = body mass index; eGFR = estimated glomerular filtration rate; FDR = false discovery rate; GFAP = glial fibrillary acidic protein; LMN = lower motor neuron; N4PD = Neurology 4-Plex D; NfL = neurofilament light chain; p-tau = phosphorylated tau.

**eTable 6:**
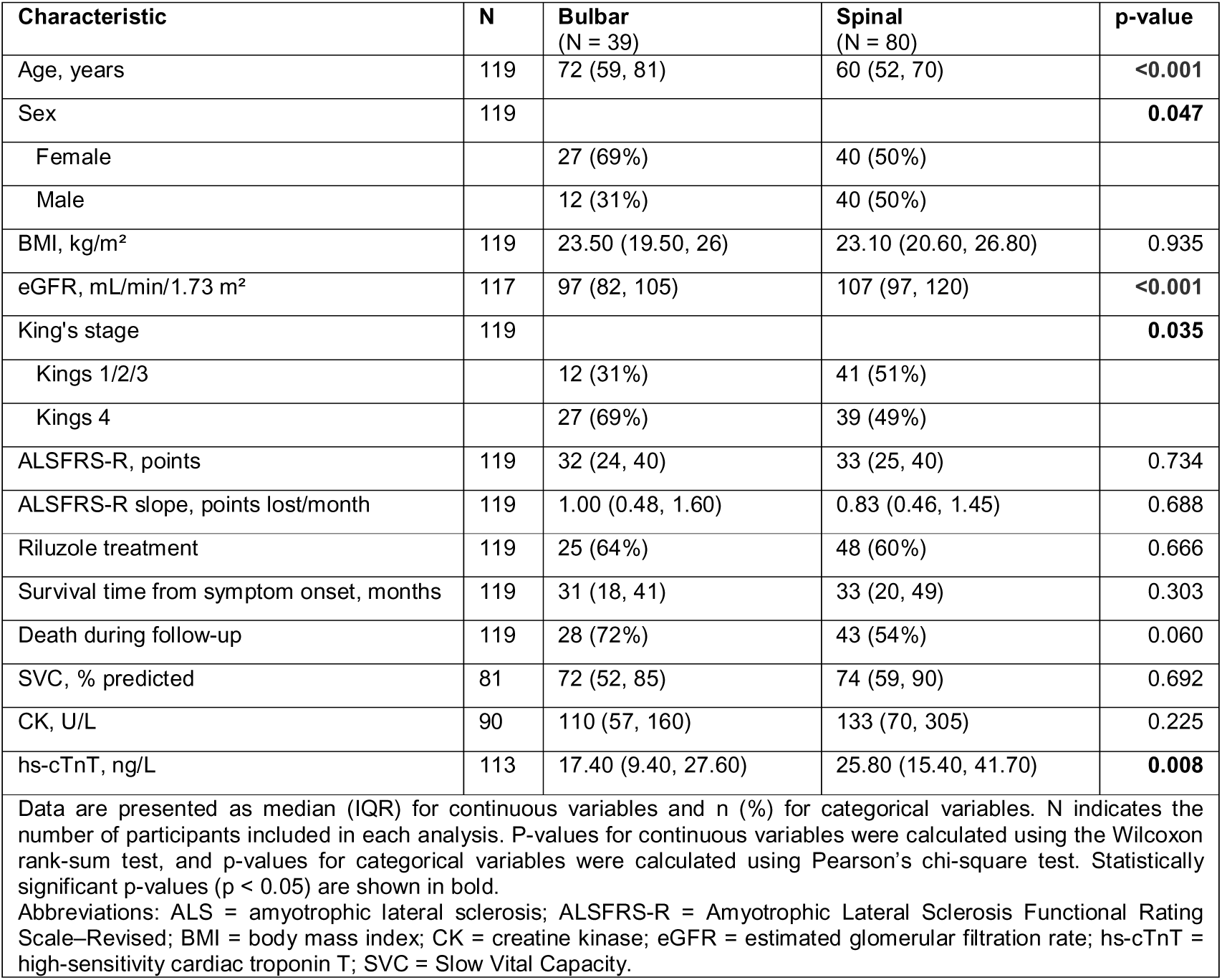
Demographics by ALS form of onset.

| Characteristic | N | Bulbar<br>(N = 39) | Spinal<br>(N = 80) | p-value |
| --- | --- | --- | --- | --- |
| Age, years | 119 | 72 (59, 81) | 60 (52, 70) | **<0.001** |
| Sex | 119 |  |  | **0.047** |
| Female |  | 27 (69%) | 40 (50%) |  |
| Male |  | 12 (31%) | 40 (50%) |  |
| BMI, kg/m<sup>2</sup> | 119 | 23.50 (19.50, 26) | 23.10 (20.60, 26.80) | 0.935 |
| eGFR, mL/min/1.73 m<sup>2</sup> | 117 | 97 (82, 105) | 107 (97, 120) | **<0.001** |
| King's stage | 119 |  |  | **0.035** |
| Kings 1/2/3 |  | 12 (31%) | 41 (51%) |  |
| Kings 4 |  | 27 (69%) | 39 (49%) |  |
| ALSFRS-R, points | 119 | 32 (24, 40) | 33 (25, 40) | 0.734 |
| ALSFRS-R slope, points lost/month | 119 | 1.00 (0.48, 1.60) | 0.83 (0.46, 1.45) | 0.688 |
| Riluzole treatment | 119 | 25 (64%) | 48 (60%) | 0.666 |
| Survival time from symptom onset, months | 119 | 31 (18, 41) | 33 (20, 49) | 0.303 |
| Death during follow-up | 119 | 28 (72%) | 43 (54%) | 0.060 |
| SVC, % predicted | 81 | 72 (52, 85) | 74 (59, 90) | 0.692 |
| CK, U/L | 90 | 110 (57, 160) | 133 (70, 305) | 0.225 |
| hs-cTnT, ng/L | 113 | 17.40 (9.40, 27.60) | 25.80 (15.40, 41.70) | **0.008** |
| Data are presented as median (IQR) for continuous variables and n (%) for categorical variables. N indicates the number of participants included in each analysis. P-values for continuous variables were calculated using the Wilcoxon rank-sum test, and p-values for categorical variables were calculated using Pearson's chi-square test. Statistically significant p-values (p < 0.05) are shown in bold.<br>Abbreviations: ALS = amyotrophic lateral sclerosis; ALSFRS-R = Amyotrophic Lateral Sclerosis Functional Rating Scale-Revised; BMI = body mass index; CK = creatine kinase; eGFR = estimated glomerular filtration rate; hs-cTnT = high-sensitivity cardiac troponin T; SVC = Slow Vital Capacity. |  |  |  |  |

**eTable 7:** Associations between ALS form of onset and blood-based biomarkers.

| Biomarker | Bulbar<br>(N=39) | Spinal<br>(N=80) | Model 1 |  | Model 2 |  | Model 3 |  |  |
| --- | --- | --- | --- | --- | --- | --- | --- | --- | --- |
|  |  |  | N | p-value | N | p-value | N | p-value | FDR<br>q-value |
| Plasma NfL (Lumipulse) | 105.70 (74, 153.34) | 90.82 (51.49, 153.20) | 119 | 0.066 | 117 | 0.440 | 117 | 0.551 | 0.587 |
| Serum NfL (Lumipulse) | 116.26 (83.20, 153.29) | 88.16 (49.78, 148.51) | 119 | **0.017** | 117 | 0.166 | 117 | 0.230 | 0.438 |
| Plasma NfL (singleplex, Simoa) | 73 (46.95, 93.35) | 58.1 (34.50, 89.38) | 119 | **0.031** | 117 | 0.199 | 117 | 0.274 | 0.438 |
| Serum NfL (singleplex, Simoa) | 97.10 (68.65, 123.50) | 76.20 (45.40, 119) | 119 | **0.018** | 117 | 0.172 | 117 | 0.248 | 0.438 |
| Plasma NfL (N4PD, Simoa) | 67.57 (48.67, 91.52) | 59.71 (33.32, 84.96) | 119 | **0.020** | 117 | 0.152 | 117 | 0.207 | 0.438 |
| Serum NfL (N4PD, Simoa) | 90.88 (65.58, 128.31) | 74.61 (46.20, 116.43) | 119 | **0.019** | 117 | 0.178 | 117 | 0.247 | 0.438 |
| Plasma p-tau181 (Simoa) | 40.19 (23.02, 62.75) | 46.10 (28.54, 88.61) | 118 | 0.154 | 116 | 0.300 | 116 | 0.234 | 0.438 |
| Serum p-tau181 (Simoa) | 34.00 (16.30, 46.15) | 36.90 (16.80, 59.70) | 116 | 0.426 | 114 | 0.446 | 114 | 0.371 | 0.469 |
| Plasma p-tau217 (Lumipulse) | 0.18 (0.12, 0.31) | 0.21 (0.15, 0.36) | 119 | 0.076 | 117 | 0.053 | 117 | **0.022** | 0.359 |
| Serum p-tau217 (Lumipulse) | 0.20 (0.14, 0.32) | 0.22 (0.14, 0.38) | 119 | 0.241 | 117 | 0.154 | 117 | 0.076 | 0.438 |
| Plasma p-tau231 (Simoa) | 3.56 (2.29, 5.93) | 4.72 (3.13, 8.03) | 116 | 0.057 | 116 | 0.105 | 116 | 0.085 | 0.438 |
| Serum p-tau231 (Simoa) | 1.56 (0.87, 2.56) | 1.83 (1.22, 3.57) | 114 | 0.143 | 112 | 0.228 | 112 | 0.259 | 0.438 |
| Plasma BD-tau (Simoa) | 8.76 (7.00, 10.81) | 8.97 (7.14, 12.66) | 119 | 0.857 | 117 | 0.654 | 117 | 0.334 | 0.469 |
| Serum BD-tau (Simoa) | 9.92 (7.42, 13.85) | 9.73 (7.54, 14.14) | 119 | 0.645 | 117 | 0.775 | 117 | 0.498 | 0.569 |
| Plasma GFAP (Simoa) | 95.35 (67.62, 135.1) | 78 (50.30, 109.34) | 119 | **0.018** | 117 | 0.717 | 117 | 0.618 | 0.618 |
| Serum GFAP (Simoa) | 114.88 (78.30, 154) | 97.40 (57.91, 136.78) | 119 | 0.065 | 117 | 0.507 | 117 | 0.381 | 0.469 |

Data are presented as median (IQR) in pg/mL. N indicates the number of participants included in each analysis. Group comparisons were performed using analysis of covariance (ANCOVA). Model 1: unadjusted. Model 2: adjusted for age, sex, BMI and eGFR. Model 3: adjusted for age, sex, BMI, eGFR, riluzole treatment, King's stage and ALSFRS-R. FDR q-values (Benjamini-Hochberg) are shown for Model 3; p- and q-values < 0.05 shown in bold.
Abbreviations: ALSFRS-R = Amyotrophic Lateral Sclerosis Functional Rating Scale-Revised; BD-tau = brain-derived tau; BMI = body mass index; eGFR = estimated glomerular filtration rate; FDR = false discovery rate; GFAP = glial fibrillary acidic protein; N4PD = Neurology 4-Plex D; NfL = neurofilament light chain; p-tau = phosphorylated tau.

**eTable 8:** Demographics by treatment.

| Characteristic | N | Treated<br>(N = 73) | Not Treated<br>(N = 46) | p-value |
| --- | --- | --- | --- | --- |
| Age, years | 119 | 59 (52, 72) | 68 (60, 77) | **0.006** |
| Sex | 119 |  |  | **0.04** |
| Female |  | 47 (64%) | 20 (43%) |  |
| Male |  | 26 (36%) | 26 (57%) |  |
| BMI, kg/m<sup>2</sup> | 119 | 22.90 (19.60, 25) | 24.30 (21.30, 28) | 0.051 |
| eGFR, mL/min/1.73 m<sup>2</sup> | 117 | 105.98 (97.02, 118.21) | 98.32 (90.86, 109.48) | **0.006** |
| ALS onset form | 119 |  |  | 0.817 |
| Bulbar |  | 25 (34%) | 14 (30%) |  |
| Spinal |  | 48 (66%) | 32 (70%) |  |
| King's stage | 119 |  |  | 0.996 |
| Kings 1/2/3 |  | 32 (44%) | 21 (46%) |  |
| Kings 4 |  | 41 (56%) | 25 (54%) |  |
| ALSFRS-R, points | 119 | 32 (24, 40) | 34.50 (28, 40) | 0.174 |
| ALSFRS-R slope, points lost/month | 119 | 0.78 (0.42, 1.40) | 1.12 (0.56, 1.63) | 0.119 |
| Survival time from symptom onset, months | 119 | 32.65 (24.03, 54.37) | 24.55 (18.97, 35.42) | **0.01** |
| Death during follow-up | 119 | 40 (55%) | 31 (67%) | 0.241 |
| SVC, % predicted | 81 | 74 (57, 90) | 73.50 (59, 99) | 0.44 |
| CK, U/L | 90 | 98 (60, 305) | 146 (65, 229) | 0.568 |
| hs-cTnT, ng/L | 113 | 20.49 (11.59, 34.67) | 27.35 (15.40, 36.16) | 0.286 |
| Data are presented as median (IQR) for continuous variables and n (%) for categorical variables. N indicates the number of participants included in each analysis. P-values for continuous variables were calculated using the Wilcoxon rank-sum test, and p-values for categorical variables were calculated using Pearson's chi-square test. Statistically significant p-values ($p < 0.05$) are shown in bold.<br>Abbreviations: ALS = amyotrophic lateral sclerosis; ALSFRS-R = Amyotrophic Lateral Sclerosis Functional Rating Scale-Revised; BMI = body mass index; CK = creatine kinase; eGFR = estimated glomerular filtration rate; hs-cTnT = high-sensitivity cardiac troponin T; SVC = Slow Vital Capacity. |  |  |  |  |

**eTable 9:** Associations between riluzole treatment status and blood-based biomarkers.

| Biomarker | Treated<br>(N=73) | Untreated<br>(N=46) | Model 1 |  | Model 2 |  | Model 3 |  |  |
| --- | --- | --- | --- | --- | --- | --- | --- | --- | --- |
|  |  |  | N | p-value | N | p-value | N | p-value | FDR q-value |
| Plasma NfL (Lumipulse) | 90.93 (58.81, 152.97) | 96.51 (67.91, 152.52) | 119 | 0.951 | 117 | 0.786 | 117 | 0.669 | 0.969 |
| Serum NfL (Lumipulse) | 96.49 (65.57, 147.62) | 93.66 (66.72, 154.18) | 119 | 0.885 | 117 | 0.875 | 117 | 0.690 | 0.969 |
| Plasma NfL (singleplex, Simoa) | 64.40 (39.9, 87.10) | 60.55 (42.05, 94.52) | 119 | 0.889 | 117 | 0.799 | 117 | 0.627 | 0.969 |
| Serum NfL (singleplex, Simoa) | 87.80 (54.30, 117) | 80.35 (57.43, 125) | 119 | 0.803 | 117 | 0.928 | 117 | 0.756 | 0.969 |
| Plasma NfL (N4PD, Simoa) | 62.33 (38.86, 92.27) | 64.45 (44.93, 86.85) | 119 | 0.808 | 117 | 0.928 | 117 | 0.727 | 0.969 |
| Serum NfL (N4PD, Simoa) | 85.14 (51.12, 123.31) | 81.28 (53.59, 122.40) | 119 | 0.715 | 117 | 0.839 | 117 | 0.969 | 0.969 |
| Plasma p-tau181 (Simoa) | 39.99 (27.49, 77.12) | 54.59 (28.62, 93.82) | 118 | 0.273 | 116 | 0.900 | 116 | 0.902 | 0.969 |
| Serum p-tau181 (Simoa) | 31.40 (17.45, 53.8) | 38.70 (14.50, 63.30) | 116 | 0.384 | 114 | 0.562 | 114 | 0.638 | 0.969 |
| Plasma p-tau217 (Lumipulse) | 0.19 (0.14, 0.32) | 0.20 (0.15, 0.39) | 119 | 0.333 | 117 | 0.998 | 117 | 0.937 | 0.969 |
| Serum p-tau217 (Lumipulse) | 0.20 (0.14, 0.35) | 0.21 (0.16, 0.43) | 119 | 0.323 | 117 | 0.940 | 117 | 0.916 | 0.969 |
| Plasma p-tau231 (Simoa) | 4.22 (2.81, 6.85) | 4.69 (2.67, 7.9) | 116 | 0.342 | 116 | 0.808 | 116 | 0.960 | 0.969 |
| Serum p-tau231 (Simoa) | 1.76 (1.10, 3.11) | 1.71 (1.20, 3.19) | 114 | 0.405 | 112 | 0.752 | 112 | 0.808 | 0.969 |
| Plasma BD-tau (Simoa) | 8.73 (6.96, 11.23) | 9.31 (7.07, 13.23) | 119 | 0.321 | 117 | 0.744 | 117 | 0.770 | 0.969 |
| Serum BD-tau (Simoa) | 9.87 (7.73, 13.06) | 9.87 (7.16, 15.89) | 119 | 0.354 | 117 | 0.667 | 117 | 0.772 | 0.969 |
| Plasma GFAP (Simoa) | 87.29 (57.13, 118.30) | 86.62 (49.55, 120.57) | 119 | 0.646 | 117 | 0.128 | 117 | 0.188 | 0.969 |
| Serum GFAP (Simoa) | 101.06 (66.17, 138.32) | 103.82 (62.04, 154.99) | 119 | 0.719 | 117 | 0.234 | 117 | 0.282 | 0.969 |

Data are presented as median (IQR) in pg/mL. N indicates the number of participants included in each analysis. Group comparisons were performed using analysis of covariance (ANCOVA). Model 1: unadjusted. Model 2: adjusted for age, sex, BMI and eGFR. Model 3: adjusted for age, sex, BMI, eGFR, ALS onset form, King's stage and ALSFRS-R. FDR q-values (Benjamini-Hochberg) are shown for Model 3; p- and q-values < 0.05 shown in bold.
Abbreviations: ALS = amyotrophic lateral sclerosis; ALSFRS-R = Amyotrophic Lateral Sclerosis Functional Rating Scale-Revised; BD-tau = brain-derived tau; BMI = body mass index; eGFR = estimated glomerular filtration rate; FDR = false discovery rate; GFAP = glial fibrillary acidic protein; N4PD = Neurology 4-Plex D; NfL = neurofilament light chain; p-tau = phosphorylated tau.

**eTable 10:** Associations between baseline ALSFRS-R and blood-based biomarkers.

| Biomarker | Model 1 |  |  | Model 2 |  |  | Model 3 |  |  |  |
| --- | --- | --- | --- | --- | --- | --- | --- | --- | --- | --- |
| | N | $\beta$ (95% CI) | p-value | N | $\beta$ (95% CI) | p-value | N | $\beta$ (95% CI) | p-value | FDR q-value |
| Plasma NfL (Lumipulse) | 119 | -4.006 (-6.634, -1.379) | **0.003** | 117 | -3.190 (-5.824, -0.557) | **0.018** | 117 | -2.187 (-4.520, 0.146) | 0.066 | 0.085 |
| Serum NfL (Lumipulse) | 119 | -4.099 (-6.686, -1.511) | **0.002** | 117 | -3.314 (-5.910, -0.717) | **0.013** | 117 | -2.270 (-4.588, 0.047) | 0.055 | 0.085 |
| Plasma NfL (singleplex, Simoa) | 119 | -4.403 (-7.048, -1.758) | **0.001** | 117 | -3.606 (-6.244, -0.969) | **0.008** | 117 | -2.468 (-4.828, -0.107) | **0.041** | 0.072 |
| Serum NfL (singleplex, Simoa) | 119 | -4.270 (-7.000, -1.539) | **0.002** | 117 | -3.435 (-6.185, -0.685) | **0.015** | 117 | -2.273 (-4.731, 0.184) | 0.069 | 0.085 |
| Plasma NfL (N4PD, Simoa) | 119 | -4.179 (-6.967, -1.391) | **0.004** | 117 | -3.372 (-6.134, -0.611) | **0.017** | 117 | -2.373 (-4.829, 0.083) | 0.058 | 0.085 |
| Serum NfL (N4PD, Simoa) | 119 | -3.738 (-6.377, -1.098) | **0.006** | 117 | -2.974 (-5.603, -0.345) | **0.027** | 117 | -2.095 (-4.423, 0.233) | 0.077 | 0.088 |
| Plasma p-tau181 (Simoa) | 118 | -1.255 (-3.535, 1.024) | 0.278 | 116 | -2.885 (-5.196, -0.573) | **0.015** | 116 | -2.401 (-4.445, -0.357) | **0.022** | **0.047** |
| Serum p-tau181 (Simoa) | 116 | -0.669 (-2.480, 1.142) | 0.466 | 114 | -2.019 (-3.834, -0.203) | **0.030** | 114 | -1.850 (-3.445, -0.255) | **0.023** | **0.047** |
| Plasma p-tau217 (Lumipulse) | 119 | -3.493 (-6.193, -0.793) | **0.012** | 117 | -5.225 (-7.973, -2.477) | **<0.001** | 117 | -4.426 (-6.893, -1.958) | **<0.001** | **0.003** |
| Serum p-tau217 (Lumipulse) | 119 | -3.197 (-5.753, -0.641) | **0.015** | 117 | -4.985 (-7.580, -2.390) | **<0.001** | 117 | -4.060 (-6.384, -1.737) | **<0.001** | **0.003** |
| Plasma p-tau231 (Simoa) | 116 | -1.313 (-3.764, 1.139) | 0.291 | 116 | -2.221 (-4.653, 0.211) | 0.073 | 116 | -1.543 (-3.736, 0.649) | 0.166 | 0.166 |
| Serum p-tau231 (Simoa) | 114 | -0.034 (-2.209, 2.140) | 0.975 | 112 | -1.161 (-3.281, 0.959) | 0.280 | 112 | -1.491 (-3.367, 0.384) | 0.118 | 0.126 |
| Plasma BD-tau (Simoa) | 119 | -6.900 (-10.736, -3.063) | **<0.001** | 117 | -9.175 (-12.860, -5.491) | **<0.001** | 117 | -6.872 (-10.280, -3.464) | **<0.001** | **<0.001** |
| Serum BD-tau (Simoa) | 119 | -6.175 (-9.735, -2.614) | **<0.001** | 117 | -8.336 (-11.787, -4.885) | **<0.001** | 117 | -6.639 (-9.741, -3.537) | **<0.001** | **<0.001** |
| Plasma GFAP (Simoa) | 119 | -3.417 (-6.267, -0.567) | **0.019** | 117 | -4.471 (-8.330, -0.612) | **0.024** | 117 | -4.783 (-8.125, -1.441) | **0.005** | **0.015** |
| Serum GFAP (Simoa) | 119 | -3.470 (-5.999, -0.941) | **0.008** | 117 | -4.263 (-7.476, -1.049) | **0.010** | 117 | -3.991 (-6.789, -1.193) | **0.006** | **0.015** |

$\beta$ coefficients and 95% confidence intervals (CIs) are reported from linear regression models examining the association between log-transformed blood-based biomarkers and ALSFRS-R. N indicates the number of participants included in each analysis. Model 1: unadjusted. Model 2: adjusted for age, sex, BMI and eGFR. Model 3: adjusted for age, sex, BMI, eGFR, ALS onset form, King's stage and riluzole treatment. FDR q-values (Benjamini–Hochberg) are shown for Model 3; p- and q-values < 0.05 shown in bold. Abbreviations: ALS = amyotrophic lateral sclerosis; ALSFRS-R = Amyotrophic Lateral Sclerosis Functional Rating Scale–Revised; BD-tau = brain-derived tau; BMI = body mass index; eGFR = estimated glomerular filtration rate; FDR = false discovery rate; GFAP = glial fibrillary acidic protein; N4PD = Neurology 4-Plex D; NfL = neurofilament light chain; p-tau = phosphorylated tau.

**eTable 11:**
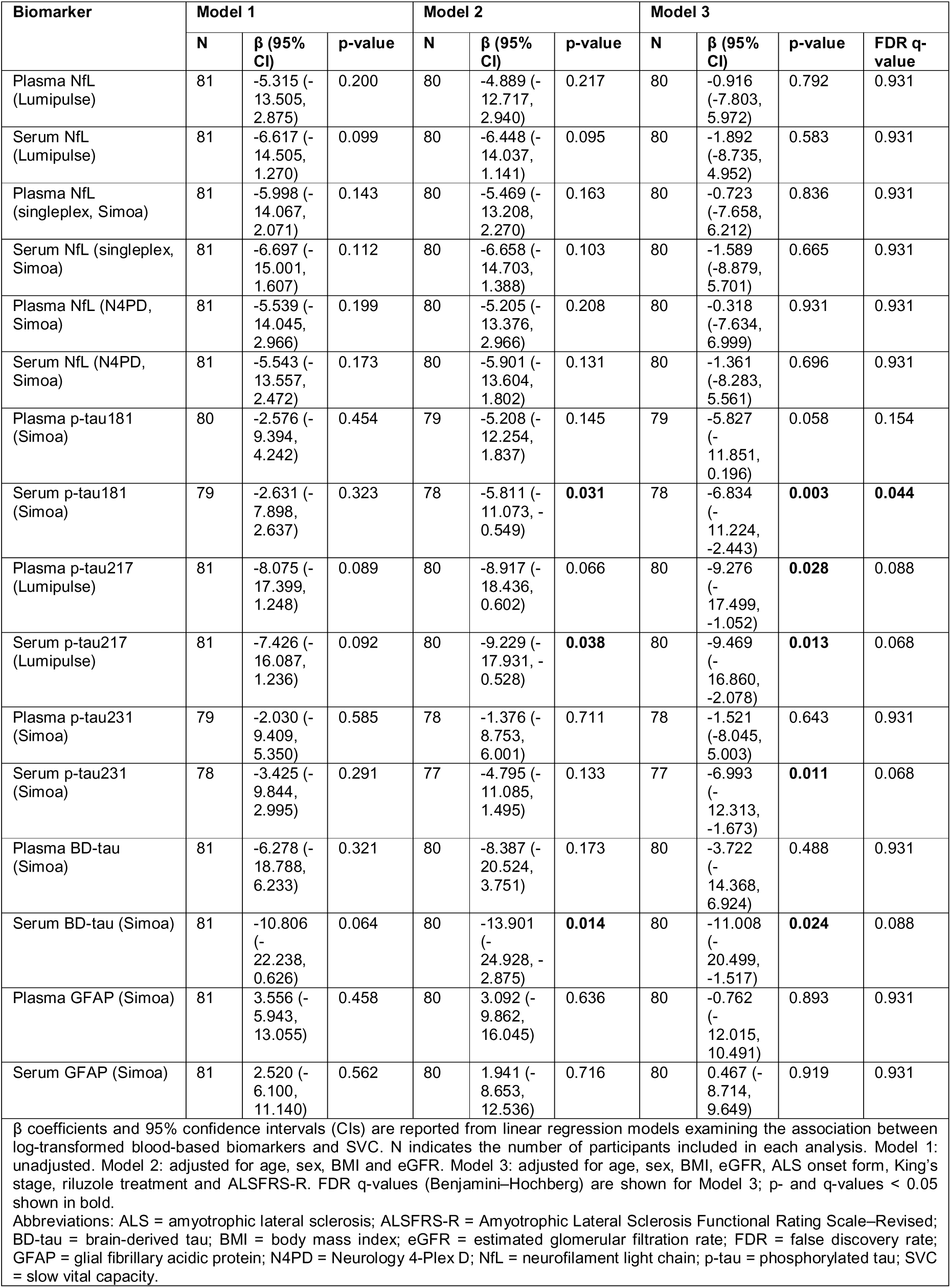
Associations between SVC and blood-based biomarkers.

| Biomarker | Model 1 |  |  | Model 2 |  |  | Model 3 |  |  |  |
| --- | --- | --- | --- | --- | --- | --- | --- | --- | --- | --- |
| | N | $\beta$ (95% CI) | p-value | N | $\beta$ (95% CI) | p-value | N | $\beta$ (95% CI) | p-value | FDR q-value |
| Plasma NfL (Lumipulse) | 81 | -5.315 (-13.505, 2.875) | 0.200 | 80 | -4.889 (-12.717, 2.940) | 0.217 | 80 | -0.916 (-7.803, 5.972) | 0.792 | 0.931 |
| Serum NfL (Lumipulse) | 81 | -6.617 (-14.505, 1.270) | 0.099 | 80 | -6.448 (-14.037, 1.141) | 0.095 | 80 | -1.892 (-8.735, 4.952) | 0.583 | 0.931 |
| Plasma NfL (singleplex, Simoa) | 81 | -5.998 (-14.067, 2.071) | 0.143 | 80 | -5.469 (-13.208, 2.270) | 0.163 | 80 | -0.723 (-7.658, 6.212) | 0.836 | 0.931 |
| Serum NfL (singleplex, Simoa) | 81 | -6.697 (-15.001, 1.607) | 0.112 | 80 | -6.658 (-14.703, 1.388) | 0.103 | 80 | -1.589 (-8.879, 5.701) | 0.665 | 0.931 |
| Plasma NfL (N4PD, Simoa) | 81 | -5.539 (-14.045, 2.966) | 0.199 | 80 | -5.205 (-13.376, 2.966) | 0.208 | 80 | -0.318 (-7.634, 6.999) | 0.931 | 0.931 |
| Serum NfL (N4PD, Simoa) | 81 | -5.543 (-13.557, 2.472) | 0.173 | 80 | -5.901 (-13.604, 1.802) | 0.131 | 80 | -1.361 (-8.283, 5.561) | 0.696 | 0.931 |
| Plasma p-tau181 (Simoa) | 80 | -2.576 (-9.394, 4.242) | 0.454 | 79 | -5.208 (-12.254, 1.837) | 0.145 | 79 | -5.827 (-11.851, 0.196) | 0.058 | 0.154 |
| Serum p-tau181 (Simoa) | 79 | -2.631 (-7.898, 2.637) | 0.323 | 78 | -5.811 (-11.073, -0.549) | **0.031** | 78 | -6.834 (-11.224, -2.443) | **0.003** | **0.044** |
| Plasma p-tau217 (Lumipulse) | 81 | -8.075 (-17.399, 1.248) | 0.089 | 80 | -8.917 (-18.436, 0.602) | 0.066 | 80 | -9.276 (-17.499, -1.052) | **0.028** | 0.088 |
| Serum p-tau217 (Lumipulse) | 81 | -7.426 (-16.087, 1.236) | 0.092 | 80 | -9.229 (-17.931, -0.528) | **0.038** | 80 | -9.469 (-16.860, -2.078) | **0.013** | 0.068 |
| Plasma p-tau231 (Simoa) | 79 | -2.030 (-9.409, 5.350) | 0.585 | 78 | -1.376 (-8.753, 6.001) | 0.711 | 78 | -1.521 (-8.045, 5.003) | 0.643 | 0.931 |
| Serum p-tau231 (Simoa) | 78 | -3.425 (-9.844, 2.995) | 0.291 | 77 | -4.795 (-11.085, 1.495) | 0.133 | 77 | -6.993 (-12.313, -1.673) | **0.011** | 0.068 |
| Plasma BD-tau (Simoa) | 81 | -6.278 (-18.788, 6.233) | 0.321 | 80 | -8.387 (-20.524, 3.751) | 0.173 | 80 | -3.722 (-14.368, 6.924) | 0.488 | 0.931 |
| Serum BD-tau (Simoa) | 81 | -10.806 (-22.238, 0.626) | 0.064 | 80 | -13.901 (-24.928, -2.875) | **0.014** | 80 | -11.008 (-20.499, -1.517) | **0.024** | 0.088 |
| Plasma GFAP (Simoa) | 81 | 3.556 (-5.943, 13.055) | 0.458 | 80 | 3.092 (-9.862, 16.045) | 0.636 | 80 | -0.762 (-12.015, 10.491) | 0.893 | 0.931 |
| Serum GFAP (Simoa) | 81 | 2.520 (-6.100, 11.140) | 0.562 | 80 | 1.941 (-8.653, 12.536) | 0.716 | 80 | 0.467 (-8.714, 9.649) | 0.919 | 0.931 |

$\beta$ coefficients and 95% confidence intervals (CIs) are reported from linear regression models examining the association between log-transformed blood-based biomarkers and SVC. N indicates the number of participants included in each analysis. Model 1: unadjusted. Model 2: adjusted for age, sex, BMI and eGFR. Model 3: adjusted for age, sex, BMI, eGFR, ALS onset form, King's stage, riluzole treatment and ALSFRS-R. FDR q-values (Benjamini-Hochberg) are shown for Model 3; p- and q-values < 0.05 shown in bold.
Abbreviations: ALS = amyotrophic lateral sclerosis; ALSFRS-R = Amyotrophic Lateral Sclerosis Functional Rating Scale-Revised; BD-tau = brain-derived tau; BMI = body mass index; eGFR = estimated glomerular filtration rate; FDR = false discovery rate; GFAP = glial fibrillary acidic protein; N4PD = Neurology 4-Plex D; NfL = neurofilament light chain; p-tau = phosphorylated tau; SVC = slow vital capacity.

**eTable 12:** Associations between Creatine Kinase and blood-based biomarkers.

| Biomarker | Model 1 |  |  | Model 2 |  |  | Model 3 |  |  |  |
| --- | --- | --- | --- | --- | --- | --- | --- | --- | --- | --- |
| | N | $\beta$ (95% CI) | p-value | N | $\beta$ (95% CI) | p-value | N | $\beta$ (95% CI) | p-value | FDR q-value |
| Plasma NfL (Lumipulse) | 90 | -0.366 (-0.680, -0.053) | **0.023** | 90 | -0.168 (-0.463, 0.126) | 0.260 | 90 | -0.127 (-0.433, 0.179) | 0.413 | 0.600 |
| Serum NfL (Lumipulse) | 90 | -0.312 (-0.622, -0.002) | **0.049** | 90 | -0.113 (-0.405, 0.180) | 0.446 | 90 | -0.062 (-0.367, 0.244) | 0.688 | 0.743 |
| Plasma NfL (singleplex, Simoa) | 90 | -0.286 (-0.609, 0.037) | 0.082 | 90 | -0.100 (-0.401, 0.201) | 0.509 | 90 | -0.047 (-0.363, 0.268) | 0.765 | 0.765 |
| Serum NfL (singleplex, Simoa) | 90 | -0.331 (-0.660, -0.001) | **0.049** | 90 | -0.135 (-0.446, 0.176) | 0.389 | 90 | -0.083 (-0.408, 0.242) | 0.614 | 0.743 |
| Plasma NfL (N4PD, Simoa) | 90 | -0.304 (-0.635, 0.026) | 0.071 | 90 | -0.125 (-0.432, 0.181) | 0.419 | 90 | -0.079 (-0.398, 0.241) | 0.624 | 0.743 |
| Serum NfL (N4PD, Simoa) | 90 | -0.287 (-0.601, 0.026) | 0.072 | 90 | -0.101 (-0.394, 0.193) | 0.497 | 90 | -0.060 (-0.363, 0.244) | 0.697 | 0.743 |
| Plasma p-tau181 (Simoa) | 89 | 0.627 (0.370, 0.885) | **<0.001** | 89 | 0.636 (0.415, 0.856) | **<0.001** | 89 | 0.704 (0.480, 0.928) | **<0.001** | **<0.001** |
| Serum p-tau181 (Simoa) | 88 | 0.468 (0.273, 0.664) | **<0.001** | 88 | 0.455 (0.281, 0.628) | **<0.001** | 88 | 0.501 (0.325, 0.677) | **<0.001** | **<0.001** |
| Plasma p-tau217 (Lumipulse) | 90 | 0.363 (0.020, 0.707) | **0.038** | 90 | 0.486 (0.179, 0.793) | **0.002** | 90 | 0.623 (0.291, 0.955) | **<0.001** | **0.001** |
| Serum p-tau217 (Lumipulse) | 90 | 0.269 (-0.058, 0.597) | 0.106 | 90 | 0.403 (0.105, 0.701) | **0.009** | 90 | 0.502 (0.184, 0.819) | **0.002** | **0.006** |
| Plasma p-tau231 (Simoa) | 88 | 0.602 (0.315, 0.889) | **<0.001** | 88 | 0.630 (0.393, 0.867) | **<0.001** | 88 | 0.699 (0.461, 0.937) | **<0.001** | **<0.001** |
| Serum p-tau231 (Simoa) | 86 | 0.625 (0.386, 0.864) | **<0.001** | 86 | 0.564 (0.357, 0.772) | **<0.001** | 86 | 0.586 (0.377, 0.795) | **<0.001** | **<0.001** |
| Plasma BD-tau (Simoa) | 90 | 0.176 (-0.287, 0.639) | 0.452 | 90 | 0.415 (-0.010, 0.840) | 0.055 | 90 | 0.644 (0.176, 1.112) | **0.008** | **0.017** |
| Serum BD-tau (Simoa) | 90 | 0.154 (-0.280, 0.587) | 0.483 | 90 | 0.375 (-0.027, 0.777) | 0.067 | 90 | 0.564 (0.120, 1.008) | **0.014** | **0.027** |
| Plasma GFAP (Simoa) | 90 | -0.591 (-0.921, -0.262) | **<0.001** | 90 | -0.230 (-0.687, 0.227) | 0.320 | 90 | -0.293 (-0.765, 0.180) | 0.222 | 0.354 |
| Serum GFAP (Simoa) | 90 | -0.525 (-0.824, -0.226) | **<0.001** | 90 | -0.220 (-0.601, 0.161) | 0.255 | 90 | -0.251 (-0.648, 0.146) | 0.212 | 0.354 |

$\beta$ coefficients and 95% confidence intervals (CIs) are reported from linear regression models examining the association between log-transformed blood-based biomarkers and creatine kinase. N indicates the number of participants included in each analysis. Model 1: unadjusted. Model 2: adjusted for age, sex, BMI and eGFR. Model 3: adjusted for age, sex, BMI, eGFR, ALS onset form, King's stage, riluzole treatment, and ALSFRS-R. FDR q-values (Benjamini–Hochberg) are shown for Model 3; p- and q-values < 0.05 shown in bold.
Abbreviations: ALS = amyotrophic lateral sclerosis; ALSFRS-R = Amyotrophic Lateral Sclerosis Functional Rating Scale–Revised; BD-tau = brain-derived tau; BMI = body mass index; eGFR = estimated glomerular filtration rate; FDR = false discovery rate; GFAP = glial fibrillary acidic protein; N4PD = Neurology 4-Plex D; NfL = neurofilament light chain; p-tau = phosphorylated tau.

**eTable 13:** Associations between hs-cTnT and blood-based biomarkers

| Biomarker | Model 1 |  |  | Model 2 |  |  | Model 3 |  |  |  |
| --- | --- | --- | --- | --- | --- | --- | --- | --- | --- | --- |
| | N | $\beta$ (95% CI) | p-value | N | $\beta$ (95% CI) | p-value | N | $\beta$ (95% CI) | p-value | FDR q-value |
| Plasma NfL (Lumipulse) | 113 | -0.138 (-0.370, 0.094) | 0.241 | 111 | -0.073 (-0.310, 0.164) | 0.541 | 111 | -0.180 (-0.395, 0.034) | 0.098 | 0.157 |
| Serum NfL (Lumipulse) | 113 | -0.147 (-0.376, 0.083) | 0.208 | 111 | -0.070 (-0.305, 0.165) | 0.558 | 111 | -0.166 (-0.381, 0.049) | 0.128 | 0.158 |
| Plasma NfL (singleplex, Simoa) | 113 | -0.149 (-0.384, 0.087) | 0.213 | 111 | -0.079 (-0.317, 0.160) | 0.515 | 111 | -0.188 (-0.406, 0.029) | 0.089 | 0.157 |
| Serum NfL (singleplex, Simoa) | 113 | -0.158 (-0.398, 0.083) | 0.196 | 111 | -0.083 (-0.329, 0.164) | 0.509 | 111 | -0.180 (-0.405, 0.044) | 0.114 | 0.158 |
| Plasma NfL (N4PD, Simoa) | 113 | -0.156 (-0.399, 0.088) | 0.209 | 111 | -0.081 (-0.327, 0.164) | 0.513 | 111 | -0.177 (-0.400, 0.047) | 0.120 | 0.158 |
| Serum NfL (N4PD, Simoa) | 113 | -0.115 (-0.347, 0.118) | 0.330 | 111 | -0.051 (-0.287, 0.184) | 0.667 | 111 | -0.137 (-0.352, 0.078) | 0.210 | 0.240 |
| Plasma p-tau181 (Simoa) | 113 | 0.618 (0.461, 0.774) | **<0.001** | 111 | 0.594 (0.422, 0.767) | **<0.001** | 111 | 0.499 (0.336, 0.662) | **<0.001** | **<0.001** |
| Serum p-tau181 (Simoa) | 112 | 0.470 (0.345, 0.595) | **<0.001** | 110 | 0.449 (0.313, 0.585) | **<0.001** | 110 | 0.379 (0.252, 0.506) | **<0.001** | **<0.001** |
| Plasma p-tau217 (Lumipulse) | 113 | 0.876 (0.707, 1.046) | **<0.001** | 111 | 0.939 (0.763, 1.116) | **<0.001** | 111 | 0.819 (0.636, 1.001) | **<0.001** | **<0.001** |
| Serum p-tau217 (Lumipulse) | 113 | 0.775 (0.608, 0.941) | **<0.001** | 111 | 0.827 (0.653, 1.002) | **<0.001** | 111 | 0.704 (0.524, 0.883) | **<0.001** | **<0.001** |
| Plasma p-tau231 (Simoa) | 111 | 0.605 (0.426, 0.784) | **<0.001** | 109 | 0.613 (0.426, 0.801) | **<0.001** | 109 | 0.515 (0.342, 0.688) | **<0.001** | **<0.001** |
| Serum p-tau231 (Simoa) | 110 | 0.498 (0.343, 0.653) | **<0.001** | 108 | 0.452 (0.292, 0.611) | **<0.001** | 108 | 0.402 (0.256, 0.548) | **<0.001** | **<0.001** |
| Plasma BD-tau (Simoa) | 111 | 1.085 (0.792, 1.379) | **<0.001** | 111 | 1.102 (0.807, 1.397) | **<0.001** | 111 | 0.901 (0.586, 1.216) | **<0.001** | **<0.001** |
| Serum BD-tau (Simoa) | 111 | 0.974 (0.717, 1.230) | **<0.001** | 111 | 1.028 (0.771, 1.284) | **<0.001** | 111 | 0.857 (0.590, 1.124) | **<0.001** | **<0.001** |
| Plasma GFAP (Simoa) | 111 | 0.076 (-0.172, 0.323) | 0.546 | 111 | 0.330 (-0.010, 0.669) | 0.057 | 111 | 0.129 (-0.194, 0.452) | 0.431 | 0.431 |
| Serum GFAP (Simoa) | 111 | 0.136 (-0.083, 0.356) | 0.221 | 111 | 0.352 (0.068, 0.635) | **0.016** | 111 | 0.141 (-0.134, 0.415) | 0.311 | 0.332 |

$\beta$ coefficients and 95% confidence intervals (CIs) are reported from linear regression models examining the association between log-transformed blood-based biomarkers and hs-cTnT. N indicates the number of participants included in each analysis. Model 1: unadjusted. Model 2: adjusted for age, sex, BMI and eGFR. Model 3: adjusted for age, sex, BMI, eGFR, ALS onset form, King's stage, riluzole treatment and ALSFRS-R. FDR q-values (Benjamini-Hochberg) are shown for Model 3; p- and q-values < 0.05 shown in bold.
Abbreviations: ALS = amyotrophic lateral sclerosis; ALSFRS-R = Amyotrophic Lateral Sclerosis Functional Rating Scale-Revised; BD-tau = brain-derived tau; BMI = body mass index; eGFR = estimated glomerular filtration rate; FDR = false discovery rate; GFAP = glial fibrillary acidic protein; hs-cTnT= high-sensitivity cardiac troponin T; N4PD = Neurology 4-Plex D; NfL = neurofilament light chain; p-tau = phosphorylated tau.

**eTable 14:** Associations between number of LMN-affected regions and blood-based biomarkers.

| Biomarker | Model 1 |  |  | Model 2 |  |  | Model 3 |  |  |  |
| --- | --- | --- | --- | --- | --- | --- | --- | --- | --- | --- |
| | N | $\beta$ (95% CI) | p-value | N | $\beta$ (95% CI) | p-value | N | $\beta$ (95% CI) | p-value | FDR q-value |
| Plasma NfL (Lumipulse) | 67 | -0.044 (-0.403, 0.315) | 0.810 | 67 | 0.116 (-0.285, 0.517) | 0.572 | 67 | -0.053 (-0.401, 0.295) | 0.766 | 0.988 |
| Serum NfL (Lumipulse) | 67 | -0.057 (-0.412, 0.298) | 0.754 | 67 | 0.108 (-0.293, 0.509) | 0.600 | 67 | -0.019 (-0.364, 0.326) | 0.914 | 0.988 |
| Plasma NfL (singleplex, Simoa) | 67 | -0.005 (-0.371, 0.361) | 0.977 | 67 | 0.154 (-0.246, 0.554) | 0.453 | 67 | -0.012 (-0.357, 0.333) | 0.946 | 0.988 |
| Serum NfL (singleplex, Simoa) | 67 | -0.023 (-0.398, 0.353) | 0.905 | 67 | 0.154 (-0.263, 0.572) | 0.472 | 67 | 0.033 (-0.326, 0.392) | 0.858 | 0.988 |
| Plasma NfL (N4PD, Simoa) | 67 | -0.025 (-0.410, 0.360) | 0.898 | 67 | 0.147 (-0.278, 0.573) | 0.500 | 67 | 0.003 (-0.363, 0.369) | 0.988 | 0.988 |
| Serum NfL (N4PD, Simoa) | 67 | -0.024 (-0.384, 0.336) | 0.897 | 67 | 0.141 (-0.261, 0.544) | 0.494 | 67 | 0.033 (-0.313, 0.379) | 0.853 | 0.988 |
| Plasma p-tau181 (Simoa) | 67 | 0.650 (0.399, 0.901) | **<0.001** | 67 | 0.688 (0.428, 0.948) | **<0.001** | 67 | 0.512 (0.251, 0.773) | **<0.001** | **0.002** |
| Serum p-tau181 (Simoa) | 65 | 0.396 (0.188, 0.604) | **<0.001** | 65 | 0.455 (0.238, 0.673) | **<0.001** | 65 | 0.318 (0.108, 0.527) | **0.004** | **0.023** |
| Plasma p-tau217 (Lumipulse) | 67 | 0.492 (0.153, 0.830) | **0.006** | 67 | 0.597 (0.230, 0.963) | **0.002** | 67 | 0.223 (-0.184, 0.629) | 0.287 | 0.766 |
| Serum p-tau217 (Lumipulse) | 67 | 0.352 (0.025, 0.678) | **0.038** | 67 | 0.475 (0.115, 0.835) | **0.012** | 67 | 0.167 (-0.212, 0.545) | 0.391 | 0.782 |
| Plasma p-tau231 (Simoa) | 65 | 0.816 (0.543, 1.090) | **<0.001** | 65 | 0.822 (0.543, 1.100) | **<0.001** | 65 | 0.615 (0.336, 0.895) | **<0.001** | **0.001** |
| Serum p-tau231 (Simoa) | 64 | 0.553 (0.277, 0.830) | **<0.001** | 64 | 0.577 (0.295, 0.858) | **<0.001** | 64 | 0.392 (0.117, 0.667) | **0.007** | **0.028** |
| Plasma BD-tau (Simoa) | 67 | 0.503 (0.049, 0.957) | **0.034** | 67 | 0.753 (0.242, 1.264) | **0.005** | 67 | 0.310 (-0.224, 0.845) | 0.260 | 0.766 |
| Serum BD-tau (Simoa) | 67 | 0.400 (-0.033, 0.832) | 0.075 | 67 | 0.643 (0.154, 1.132) | **0.012** | 67 | 0.236 (-0.276, 0.747) | 0.371 | 0.782 |
| Plasma GFAP (Simoa) | 67 | -0.085 (-0.412, 0.242) | 0.612 | 67 | 0.230 (-0.254, 0.715) | 0.355 | 67 | -0.008 (-0.443, 0.426) | 0.970 | 0.988 |
| Serum GFAP (Simoa) | 67 | -0.020 (-0.311, 0.271) | 0.892 | 67 | 0.230 (-0.163, 0.624) | 0.255 | 67 | -0.020 (-0.378, 0.337) | 0.913 | 0.988 |

$\beta$ coefficients and 95% confidence intervals (CIs) are reported from linear regression models examining the association between log-transformed blood-based biomarkers and number of LMN-affected regions. N indicates the number of participants included in each analysis. Model 1: unadjusted. Model 2: adjusted for age, sex, BMI and eGFR. Model 3: adjusted for age, sex, BMI, eGFR, ALS onset form, King's stage, riluzole treatment and ALSFRS-R. FDR q-values (Benjamini–Hochberg) are shown for Model 3; p- and q-values < 0.05 shown in bold.
Abbreviations: ALS = amyotrophic lateral sclerosis; ALSFRS-R = Amyotrophic Lateral Sclerosis Functional Rating Scale–Revised; BD-tau = brain-derived tau; BMI = body mass index; eGFR = estimated glomerular filtration rate; FDR = false discovery rate; GFAP = glial fibrillary acidic protein; LMN = lower motor neuron; N4PD = Neurology 4-Plex D; NfL = neurofilament light chain; p-tau = phosphorylated tau.

**eTable 15:** Associations between percentage of affected muscles and blood-based biomarkers.

| Biomarker | Model 1 |  |  | Model 2 |  |  | Model 3 |  |  |  |
| --- | --- | --- | --- | --- | --- | --- | --- | --- | --- | --- |
|  | N | β (95% CI) | p-value | N | β (95% CI) | p-value | N | β (95% CI) | p-value | FDR q-value |
| Plasma NfL (Lumipulse) | 63 | -0.225 (-9.352, 8.903) | 0.962 | 63 | 4.650 (-5.406, 14.706) | 0.369 | 63 | -0.111 (-8.994, 8.772) | 0.981 | 0.993 |
| Serum NfL (Lumipulse) | 63 | -1.143 (-10.161, 7.876) | 0.805 | 63 | 3.655 (-6.412, 13.722) | 0.480 | 63 | 0.042 (-8.756, 8.840) | 0.993 | 0.993 |
| Plasma NfL (singleplex, Simoa) | 63 | 0.418 (-8.872, 9.707) | 0.930 | 63 | 5.247 (-4.853, 15.347) | 0.313 | 63 | 0.237 (-8.669, 9.143) | 0.959 | 0.993 |
| Serum NfL (singleplex, Simoa) | 63 | -0.907 (-10.471, 8.657) | 0.853 | 63 | 4.040 (-6.507, 14.586) | 0.456 | 63 | 0.897 (-8.309, 10.102) | 0.849 | 0.993 |
| Plasma NfL (N4PD, Simoa) | 63 | -0.109 (-9.926, 9.708) | 0.983 | 63 | 5.006 (-5.749, 15.762) | 0.365 | 63 | 0.884 (-8.519, 10.287) | 0.855 | 0.993 |
| Serum NfL (N4PD, Simoa) | 63 | -2.286 (-11.538, 6.965) | 0.630 | 63 | 2.090 (-8.236, 12.416) | 0.693 | 63 | -0.092 (-9.079, 8.895) | 0.984 | 0.993 |
| Plasma p-tau181 (Simoa) | 63 | 14.586 (7.633, 21.540) | **<0.001** | 63 | 14.864 (7.724, 22.004) | **<0.001** | 63 | 10.888 (3.864, 17.913) | **0.004** | **0.030** |
| Serum p-tau181 (Simoa) | 61 | 8.138 (1.845, 14.431) | **0.014** | 61 | 8.774 (2.271, 15.277) | **0.011** | 61 | 6.458 (0.418, 12.498) | **0.041** | 0.164 |
| Plasma p-tau217 (Lumipulse) | 63 | 11.731 (2.793, 20.668) | **0.013** | 63 | 13.559 (4.019, 23.099) | **0.007** | 63 | 4.763 (-5.898, 15.424) | 0.385 | 0.971 |
| Serum p-tau217 (Lumipulse) | 63 | 8.844 (0.458, 17.231) | **0.043** | 63 | 11.265 (2.155, 20.374) | **0.019** | 63 | 4.728 (-4.947, 14.403) | 0.342 | 0.971 |
| Plasma p-tau231 (Simoa) | 62 | 18.560 (11.182, 25.937) | **<0.001** | 62 | 18.612 (11.299, 25.924) | **<0.001** | 62 | 13.165 (5.846, 20.485) | **<0.001** | **0.014** |
| Serum p-tau231 (Simoa) | 60 | 13.058 (5.307, 20.808) | **0.002** | 60 | 13.092 (5.285, 20.899) | **0.002** | 60 | 9.196 (1.746, 16.647) | **0.019** | 0.103 |
| Plasma BD-tau (Simoa) | 63 | 9.650 (-2.460, 21.761) | 0.124 | 63 | 14.878 (1.329, 28.427) | **0.036** | 63 | 2.465 (-12.108, 17.037) | 0.742 | 0.993 |
| Serum BD-tau (Simoa) | 63 | 5.152 (-7.060, 17.364) | 0.412 | 63 | 9.627 (-4.087, 23.341) | 0.174 | 63 | -2.065 (-16.862, 12.731) | 0.785 | 0.993 |
| Plasma GFAP (Simoa) | 63 | -3.929 (-13.138, 5.280) | 0.406 | 63 | 5.814 (-8.636, 20.263) | 0.434 | 63 | 4.073 (-8.219, 16.365) | 0.519 | 0.993 |
| Serum GFAP (Simoa) | 63 | -1.511 (-10.193, 7.171) | 0.734 | 63 | 7.710 (-4.661, 20.080) | 0.227 | 63 | 4.373 (-6.288, 15.035) | 0.425 | 0.971 |

β coefficients and 95% confidence intervals (CIs) are reported from linear regression models examining the association between log-transformed blood-based biomarkers and the percentage of affected muscles. N indicates the number of participants included in each analysis. Model 1: unadjusted. Model 2: adjusted for age, sex, BMI and eGFR. Model 3: adjusted for age, sex, BMI, eGFR, ALS onset form, King's stage, riluzole treatment and ALSFRS-R. FDR q-values (Benjamini–Hochberg) are shown for Model 3; p- and q-values < 0.05 shown in bold.
Abbreviations: ALS = amyotrophic lateral sclerosis; ALSFRS-R = Amyotrophic Lateral Sclerosis Functional Rating Scale–Revised; BD-tau = brain-derived tau; BMI = body mass index; eGFR = estimated glomerular filtration rate; FDR = false discovery rate; GFAP = glial fibrillary acidic protein; N4PD = Neurology 4-Plex D; NfL = neurofilament light chain; p-tau = phosphorylated tau.

**eTable 16:** Associations between ALSFRS-R slope and blood-based biomarkers.

| Biomarker | Model 1 |  |  | Model 2 |  |  | Model 3 |  |  |  |
| --- | --- | --- | --- | --- | --- | --- | --- | --- | --- | --- |
| | N | $\beta$ (95% CI) | p-value | N | $\beta$ (95% CI) | p-value | N | $\beta$ (95% CI) | p-value | FDR q-value |
| Plasma NfL (Lumipulse) | 119 | 0.635 (0.400, 0.869) | **<0.001** | 117 | 0.645 (0.390, 0.901) | **<0.001** | 117 | 0.509 (0.271, 0.747) | **<0.001** | **<0.001** |
| Serum NfL (Lumipulse) | 119 | 0.605 (0.372, 0.839) | **<0.001** | 117 | 0.616 (0.362, 0.870) | **<0.001** | 117 | 0.482 (0.243, 0.721) | **<0.001** | **<0.001** |
| Plasma NfL (singleplex, Simoa) | 119 | 0.641 (0.403, 0.879) | **<0.001** | 117 | 0.652 (0.395, 0.909) | **<0.001** | 117 | 0.505 (0.261, 0.749) | **<0.001** | **<0.001** |
| Serum NfL (singleplex, Simoa) | 119 | 0.589 (0.339, 0.840) | **<0.001** | 117 | 0.599 (0.326, 0.873) | **<0.001** | 117 | 0.456 (0.200, 0.713) | **<0.001** | **<0.001** |
| Plasma NfL (N4PD, Simoa) | 119 | 0.681 (0.433, 0.929) | **<0.001** | 117 | 0.691 (0.425, 0.958) | **<0.001** | 117 | 0.561 (0.312, 0.811) | **<0.001** | **<0.001** |
| Serum NfL (N4PD, Simoa) | 119 | 0.580 (0.341, 0.819) | **<0.001** | 117 | 0.585 (0.325, 0.844) | **<0.001** | 117 | 0.472 (0.233, 0.712) | **<0.001** | **<0.001** |
| Plasma p-tau181 (Simoa) | 118 | -0.037 (-0.258, 0.184) | 0.739 | 116 | -0.008 (-0.260, 0.244) | 0.949 | 116 | -0.150 (-0.378, 0.079) | 0.197 | 0.337 |
| Serum p-tau181 (Simoa) | 116 | -0.043 (-0.219, 0.132) | 0.625 | 114 | -0.034 (-0.231, 0.164) | 0.736 | 114 | -0.119 (-0.297, 0.060) | 0.190 | 0.337 |
| Plasma p-tau217 (Lumipulse) | 119 | 0.054 (-0.212, 0.320) | 0.687 | 117 | 0.092 (-0.213, 0.397) | 0.553 | 117 | -0.178 (-0.470, 0.115) | 0.232 | 0.337 |
| Serum p-tau217 (Lumipulse) | 119 | 0.051 (-0.200, 0.303) | 0.688 | 117 | 0.084 (-0.205, 0.372) | 0.567 | 117 | -0.171 (-0.445, 0.103) | 0.219 | 0.337 |
| Plasma p-tau231 (Simoa) | 116 | -0.033 (-0.276, 0.210) | 0.789 | 114 | 0.002 (-0.270, 0.273) | 0.990 | 114 | -0.127 (-0.372, 0.119) | 0.309 | 0.411 |
| Serum p-tau231 (Simoa) | 114 | -0.100 (-0.277, 0.076) | 0.263 | 112 | -0.085 (-0.276, 0.107) | 0.383 | 112 | -0.115 (-0.291, 0.061) | 0.197 | 0.337 |
| Plasma BD-tau (Simoa) | 119 | 0.465 (0.087, 0.843) | **0.016** | 117 | 0.520 (0.105, 0.934) | **0.014** | 117 | 0.109 (-0.309, 0.527) | 0.606 | 0.746 |
| Serum BD-tau (Simoa) | 119 | 0.362 (0.010, 0.714) | **0.044** | 117 | 0.402 (0.012, 0.792) | **0.043** | 117 | 0.036 (-0.351, 0.423) | 0.854 | 0.911 |
| Plasma GFAP (Simoa) | 119 | 0.152 (-0.127, 0.430) | 0.283 | 117 | 0.114 (-0.299, 0.527) | 0.587 | 117 | -0.001 (-0.384, 0.384) | 1.000 | 1.000 |
| Serum GFAP (Simoa) | 119 | 0.127 (-0.122, 0.376) | 0.314 | 117 | 0.085 (-0.262, 0.431) | 0.629 | 117 | -0.062 (-0.383, 0.259) | 0.702 | 0.802 |

$\beta$ coefficients and 95% confidence intervals (CIs) are reported from linear regression models examining the association between log-transformed blood-based biomarkers and the ALSFRS-R slope. N indicates the number of participants included in each analysis. Model 1: unadjusted. Model 2: adjusted for age, sex, BMI and eGFR. Model 3: adjusted for age, sex, BMI, eGFR, ALS onset form, King's stage, riluzole treatment and ALSFRS-R. FDR q-values (Benjamini–Hochberg) are shown for Model 3; p- and q-values < 0.05 shown in bold.
Abbreviations: ALS = amyotrophic lateral sclerosis; ALSFRS-R = Amyotrophic Lateral Sclerosis Functional Rating Scale–Revised; BD-tau = brain-derived tau; BMI = body mass index; eGFR = estimated glomerular filtration rate; FDR = false discovery rate; GFAP = glial fibrillary acidic protein; N4PD = Neurology 4-Plex D; NfL = neurofilament light chain; p-tau = phosphorylated tau.

**eTable 17:** Cox PH regression Hazard Ratios.

| Biomarker | Model 1 |  |  | Model 2 |  |  | Model 3 |  |  |  | Model 4 |  |  |  |
| --- | --- | --- | --- | --- | --- | --- | --- | --- | --- | --- | --- | --- | --- | --- |
|  | N | HR (95% CI) | p-value | N | HR (95% CI) | p-value | N | HR (95% CI) | p-value | FDR q-value | N | HR (95% CI) | p-value | FDR q-value |
| Plasma NfL (Lumipulse) | 54 | 1.98 (1.47–2.66) | **<0.001** | 54 | 2.01 (1.45–2.78) | **<0.001** | 54 | 1.96 (1.41–2.74) | **<0.001** | **<0.001** | 33 | 2.22 (1.26–3.91) | **0.006** | 0.053 |
| Serum NfL (Lumipulse) | 54 | 1.94 (1.44–2.62) | **<0.001** | 54 | 1.94 (1.41–2.69) | **<0.001** | 54 | 1.96 (1.40–2.73) | **<0.001** | **<0.001** | 33 | 1.91 (1.12–3.26) | **0.017** | 0.053 |
| Plasma NfL (singleplex, Simoa) | 54 | 1.84 (1.39–2.44) | **<0.001** | 54 | 1.95 (1.42–2.68) | **<0.001** | 54 | 1.90 (1.37–2.63) | **<0.001** | **<0.001** | 33 | 2.06 (1.20–3.56) | **0.009** | 0.053 |
| Serum NfL (singleplex, Simoa) | 54 | 1.83 (1.36–2.47) | **<0.001** | 54 | 1.85 (1.34–2.56) | **<0.001** | 54 | 1.86 (1.34–2.59) | **<0.001** | **<0.001** | 33 | 1.92 (1.11–3.33) | **0.020** | 0.053 |
| Plasma NfL (N4PD, Simoa) | 54 | 1.83 (1.39–2.41) | **<0.001** | 54 | 1.99 (1.45–2.72) | **<0.001** | 54 | 1.97 (1.42–2.73) | **<0.001** | **<0.001** | 33 | 2.11 (1.18–3.78) | **0.012** | 0.053 |
| Serum NfL (N4PD, Simoa) | 54 | 1.91 (1.41–2.60) | **<0.001** | 54 | 1.92 (1.38–2.67) | **<0.001** | 54 | 1.95 (1.39–2.74) | **<0.001** | **<0.001** | 33 | 1.99 (1.13–3.50) | **0.017** | 0.053 |
| Plasma p-tau181 (Simoa) | 54 | 1.11 (0.85–1.44) | 0.440 | 54 | 1.22 (0.88–1.70) | 0.228 | 54 | 1.27 (0.90–1.79) | 0.168 | 0.244 | 33 | 0.99 (0.61–1.62) | 0.980 | 0.992 |
| Serum p-tau181 (Simoa) | 53 | 1.18 (0.88–1.58) | 0.275 | 53 | 1.42 (0.99–2.05) | 0.060 | 53 | 1.54 (1.07–2.22) | **0.021** | **0.049** | 32 | 1.45 (0.83–2.54) | 0.196 | 0.348 |
| Plasma p-tau217 (Lumipulse) | 54 | 1.19 (0.91–1.55) | 0.203 | 54 | 1.26 (0.90–1.76) | 0.186 | 54 | 1.25 (0.88–1.78) | 0.207 | 0.276 | 33 | 1.41 (0.79–2.51) | 0.248 | 0.377 |
| Serum p-tau217 (Lumipulse) | 54 | 1.23 (0.95–1.59) | 0.120 | 54 | 1.30 (0.95–1.77) | 0.100 | 54 | 1.32 (0.94–1.84) | 0.108 | 0.173 | 33 | 1.49 (0.87–2.57) | 0.146 | 0.292 |
| Plasma p-tau231 (Simoa) | 52 | 1.07 (0.82–1.41) | 0.604 | 52 | 1.14 (0.84–1.56) | 0.404 | 52 | 1.16 (0.83–1.64) | 0.388 | 0.477 | 32 | 1.00 (0.62–1.63) | 0.992 | 0.992 |
| Serum p-tau231 (Simoa) | 51 | 1.14 (0.86–1.51) | 0.375 | 51 | 1.33 (0.96–1.85) | 0.086 | 51 | 1.45 (1.04–2.02) | **0.029** | 0.052 | 31 | 1.36 (0.80–2.31) | 0.259 | 0.377 |
| Plasma BD-tau (Simoa) | 54 | 1.27 (0.99–1.64) | **0.065** | 54 | 1.12 (0.87–1.46) | 0.385 | 54 | 1.10 (0.82–1.48) | 0.514 | 0.588 | 33 | 1.03 (0.69–1.54) | 0.876 | 0.992 |
| Serum BD-tau (Simoa) | 54 | 1.52 (1.11–2.09) | **0.009** | 54 | 1.57 (1.08–2.29) | **0.017** | 54 | 1.55 (1.05–2.27) | **0.026** | 0.051 | 33 | 1.83 (1.02–3.26) | **0.041** | 0.094 |
| Plasma GFAP (Simoa) | 54 | 1.39 (1.05–1.83) | **0.021** | 54 | 1.03 (0.66–1.60) | 0.892 | 54 | 1.13 (0.71–1.80) | 0.600 | 0.640 | 33 | 1.22 (0.70–2.14) | 0.485 | 0.597 |
| Serum GFAP (Simoa) | 54 | 1.42 (1.06–1.89) | **0.018** | 54 | 1.13 (0.76–1.69) | 0.549 | 54 | 1.10 (0.72–1.68) | 0.655 | 0.655 | 33 | 1.24 (0.78–1.98) | 0.358 | 0.478 |

Hazard ratios (HRs) and 95% confidence intervals (CIs) are reported from Cox proportional hazards (PH) regression models using log-transformed standardized (z-score) blood-based biomarkers as predictors, and using the whole cohort as a reference group. N indicates the number of participants included in each analysis. Model 1: unadjusted. Model 2: adjusted for age, sex, BMI and eGFR. Model 3: adjusted for age, sex, BMI, eGFR, ALS onset form, riluzole treatment, Kings stage and ALSFRS-R. Model 4: adjusted for age, sex, BMI, eGFR, ALS onset form, King's stage, riluzole treatment, ALSFRS-R and the number of LMN-affected regions. FDR q-values (Benjamini–Hochberg) are shown for Models 3 and 4; p- and q-values < 0.05 shown in bold. Abbreviations: ALS = amyotrophic lateral sclerosis; ALSFRS-R = Amyotrophic Lateral Sclerosis Functional Rating Scale–Revised; BD-tau = brain-derived tau; BMI = body mass index; eGFR = estimated glomerular filtration rate; FDR = false discovery rate; GFAP = glial fibrillary acidic protein; LMN = lower motor neuron; N4PD = Neurology 4-Plex D; NfL = neurofilament light chain; p-tau = phosphorylated tau.

